# Molecular and micro-architectural mapping of gray matter alterations in psychosis

**DOI:** 10.1101/2023.12.11.23299652

**Authors:** Natalia García-San-Martín, Richard AI Bethlehem, Agoston Mihalik, Jakob Seidlitz, Isaac Sebenius, Claudio Alemán-Morillo, Lena Dorfschmidt, Golia Shafiei, Víctor Ortiz-García de la Foz, Kate Merritt, Anthony David, Sarah E. Morgan, Miguel Ruiz-Veguilla, Rosa Ayesa-Arriola, Javier Vázquez-Bourgon, Aaron Alexander-Bloch, Bratislav Misic, Edward T. Bullmore, John Suckling, Benedicto Crespo-Facorro, Lifespan Brain Chart Consortium, Rafael Romero-García

**Affiliations:** Department of Medical Physiology and Biophysics, University of Seville; Seville, Spain; Department of Psychology, University of Cambridge; Cambridge, UK; Department of Psychiatry, University of Cambridge; Cambridge, UK; Department of Child and Adolescent Psychiatry and Behavioral Science, The Children’s Hospital of Philadelphia; Philadelphia, PA, USA; Lifespan Brain Institute, The Children’s Hospital of Philadelphia and Penn Medicine; Philadelphia, PA, USA; Department of Psychiatry, Perelman School of Medicine, University of Pennsylvania; Philadelphia, Pennsylvania, USA; Department of Psychiatry. Marqués de Valdecilla University Hospital, IDIVAL, School of Medicine, University of Cantabria; Santander, Spain; Division of Psychiatry, Institute of Mental Health, UCL; London, UK; Mental Health Service, Virgen del Rocío University Hospital; Seville, Spain; Instituto de Biomedicina de Sevilla (IBiS) HUVR/CSIC/University of Seville / CIBERSAM, ISCIII; Seville, Spain; Montreal Neurological Institute, McGill University; Montreal, Canada

**Author notes:** Correspondence to: Rafael Romero García, Avda. Doctor Fedriani S/N, 41009, Seville (Spain).

## Abstract

The psychosis spectrum encompasses a heterogeneous range of clinical conditions associated with abnormal brain development. Detecting patterns of atypical neuroanatomical maturation across psychiatric disorders requires an interpretable metric standardized by age-, sex- and site-effect. The molecular and micro-architectural attributes that account for these deviations in brain structure from typical neurodevelopment are still unknown. Here, we aggregate structural magnetic resonance imaging data from 38,696 healthy controls (HC) and 1,256 psychosis-related conditions, including first-degree relatives of schizophrenia (SCZ) and schizoaffective disorder (SAD) patients (*n* = 160), individuals who had psychotic experiences (*n* = 157), patients who experienced a first episode of psychosis (FEP, *n* = 352), and individuals with chronic SCZ or SAD (*n* = 587). Using a normative modeling approach, we generated centile scores for cortical gray matter (GM) phenotypes, identifying deviations in regional volumes below the expected trajectory for all conditions, with a greater impact on the clinically diagnosed ones, FEP and chronic. Additionally, we mapped 46 neurobiological features from healthy individuals (including neurotransmitters, cell types, layer thickness, microstructure, cortical expansion, and metabolism) to these abnormal centiles using a multivariate approach. Results revealed that neurobiological features were highly co-localized with centile deviations, where metabolism (e.g., cerebral metabolic rate of oxygen (CMRGlu) and cerebral blood flow (CBF)) and neurotransmitter concentrations (e.g., serotonin (5-HT) and acetylcholine (α_4_β_2_) receptors) showed the most consistent spatial overlap with abnormal GM trajectories. Taken together these findings shed light on the vulnerability factors that may underlie atypical brain maturation during different stages of psychosis.

## Introduction

The psychosis spectrum comprises a range of psychotic disorders characterized by shared patterns of atrophy and microstructural changes/alterations in gray matter (GM) [1]. This hypothesis of a continuum spectrum is frequently supported by common symptomatology and biomarkers across different diseases, such as schizophrenia (SCZ) and schizoaffective disorder (SAD) [2]. SCZ is a severe and chronic psychotic disorder characterized by delusions and hallucinations [3] that has been associated with several risk factors, such as genetic predisposition, substance abuse, and perinatal and early environmental adversities [4]. Family history is an influential vulnerability factor with an estimated heritability of nearly 80% [5]. Accordingly, schizotypal personality disorder, a non-psychotic disorder with schizophrenic traits, is more prevalent among relatives of individuals with SCZ compared to relatives of controls [6]. The presence of subtle cognitive and behavioral abnormalities, as well as a range of cognitive impairments similar to those observed among SCZ patients, has been consistently documented in these non-affected SCZ relatives [7]. In terms of brain structure across different stages of the disorder, individuals with SCZ have shown progressive reductions in cortical GM volume [8, 9]. The initial phases of psychosis have also been extensively linked to GM changes in specific regions, and the transition to psychosis is further characterized by a progressive loss of volume [10]. Even unaffected first-degree relatives of SCZ patients have shown GM alterations, revealing that genetic factors may play an important role in abnormal brain structure [11].

In addition to SCZ, several clinical diagnostic categories are marked by psychotic symptoms, while the relationship and boundaries among them are still a matter of debate [12]. For instance, SAD is a condition characterized by the co-occurrence of schizophrenic symptoms with affective disturbance [3]. It is frequently regarded as a heterogeneous spectrum disorder, with some patients leaning more towards SCZ and others more towards affective disorders. Therefore, there is considerable overlap between SCZ and SAD in terms of symptomatology and treatment [3], showing widespread and overlapping areas of significant GM volume reduction [13]. Nevertheless, the underlying disease mechanism of these psychosis-related GM reductions remains unknown, and the relationship between neurobiology and changes in cortical structure is still unclear [7].

GM reduction appears to be linked to the pathophysiology of psychosis spectrum onset, while changes may stabilize in latter stages [14]. Beyond these age-dependent alterations in brain structure, consistent evidence also reveals sex as a differentiating factor [15]. This interplay of age- and sex-dependent neurodevelopment with the different phases of psychosis spectrum plays a crucial role in the neurodevelopmental hypothesis of psychosis [16, 17]. This hypothesis posits that genetic and environmental alterations in early (perinatal) brain development and adolescence lead to the emergence of psychotic symptomatology. Hence, understanding this complex dynamic requires of an analytical approach capable of handling maturational brain models. Identifying these deviations in clinical measurements from expected normative values can be achieved using ranked centile scores (e.g., assessment of bone strength [18], metabolic rate [19], or height and weight measurements commonly used in routine pediatric care). This strategy has been recently extended by Bethlehem *et al*. [20] which provides reference charts of brain volume to compute individual volumetric centiles normalized by age-, sex- and, importantly, site-effects. Consequently, centile scores provide a standardized and interpretable metric for detecting alterations in regional volumes, thereby contributing to identify patterns of atypical neuroanatomical maturation across psychiatric disorders.

Psychosis has been associated with specific neurobiological alterations, such as neurotransmission [21], metabolism [22], cell type [23], and microstructure [24]. Studies on prevalent genetic variations linked to SCZ have consistently pointed towards synaptic function as key factor in terms of disease risk. Specifically, dysregulation of dopaminergic neurotransmission has been detected in SCZ [3], and several neurochemical systems have been suggested to contribute to psychosis pathophysiology, including the glutamate, gamma-aminobutyric acid (GABA), serotonin, and acetylcholine neurotransmitters [21]. Complementing the role of neurotransmitters, metabolic and cellular alterations have also been associated with psychosis. Energy metabolism interacts with the disrupted balance of excitatory and inhibitory neurons in SCZ, which is maintained by glutamatergic and GABAergic signaling [22]; and SCZ patients have demonstrated abnormalities in astroglial and oligodendroglial cells [23]. Regarding microstructure, several abnormalities have been detected in SCZ patients related to myelination, neuropil organization, and expression of proteins that support neurite and synaptic integrity [24]. Co-localizing (i.e., being located in the same region of the cortex) these distinctive neurobiological features and the structural brain differences related to stages of psychosis may help understand the common and differential mechanisms involved in the ontology of this disease.

The spatial topography of volume alterations related to psychosis is not uniform across the cortex [25]; instead, certain regions appear to be more susceptible to disease pathology. This observation aligns with the regional vulnerability hypothesis, which posits that local features such as cellular composition [23], neurotransmitter receptors [21], glutamatergic metabolism [22], and gene expression [26] may play a crucial role in the pathophysiology and symptomatology of psychiatric conditions [27]. Within this context of localized susceptibility, recent research has explored the role that brain network dynamics plays in the progression of glioblastoma multiforme [28]; or the influence of network connectivity on vulnerability, resilience, and expression to the onset of SCZ [29]. For example, excessive glutamatergic neurotransmission could lead to excitotoxic cellular damage and death, potentially resulting in GM volume reduction [30]. Nevertheless, the understanding of the vulnerabilities that lead to volumetric changes is still limited.

In the present study, we aimed to characterize the molecular and micro-architectural attributes (collectively referred here as neurobiological features) that underlie the pattern of cortical atypical maturation in different psychosis-related groups. We first computed centiles from regional volumes for each group, anticipating that, in line with the neurodevelopmental hypothesis of psychosis, these scores would reveal a pattern of abnormal deviations from normative trajectories. Next, we investigated the regional vulnerability to psychosis through a multivariate model for each condition, where centiles were mapped by 46 different neurobiological features, including neurotransmitters, cell types, layer thickness, microstructure, cortical expansion, and metabolism. Finally, we explored the associations between the inter-regional vulnerability to psychosis across conditions and the neurobiological similarities.

## Subjects and Methods

### Subjects

Magnetic Resonance Imaging (MRI) data were analyzed from eight psychosis-related diagnoses, which were clustered into four groups according to their clinical profile. MRI from individuals with chronic schizophrenia (SCZ; *n* = 525; *age* = 37.63 ± 12.06) and schizoaffective disorder (SAD; *n* = 62; *age* = 33.77 ± 10.78), clustered under SCZ and SAD-chronic group, were obtained from Adolescent Brain Cognitive Development (ABCD) [31], Australian Schizophrenia Research Bank (ASRB) [32], Bipolar-Schizophrenia Network on Intermediate Phenotypes (B-SNIP) [33], UCLA Consortium for Neuropsychiatric Phenomics (CNP) LA5c Study [34], Mental Illness and Neuroscience Discovery (MIND) Institute Clinical Imaging Consortium (MCIC) [35], and UK Biobank (UKB) [36]. To be included in the chronic group, patients had to meet DSM-IV diagnostic criteria for schizophrenia or schizoaffective disorder, which require the presence of at least two psychotic symptoms during a 1-month period. Healthy controls (*n* = 38,232; *age* = 51.91 ± 23.68) comprised individuals from previous datasets without any current or previous psychotic disorder.

MRI data from individuals who were determined to have experienced a first episode of psychosis (FEP; *n* = 352; *age* = 31.43 ± 8.78), along with their respective HC (*n* = 195; *age* = 30.64 ± 7.66), were obtained from *Programa de Atención a las Fases Iniciales de Psicosis* (PAFIP) [39]. Scanning was performed prior to the initiation of any antipsychotic medication. All diagnoses were made by an experienced psychiatrist using the Structured Clinical Interview for DSM-IV (SCID-I) after 6 months of the baseline visit, confirming the presence of schizophrenia or other psychotic disorder.

Individuals who were suspected of having had psychotic experiences (PEs; *PE-suspected*; *n* = 48; *age* = 21.55 ± 1.06), or were rated as definitely having PEs (*PE-definite*; *n* = 73; *age* = 21.75 ± 1.59), or have suffered PEs with social decline and/or help-seeking (*PE-clinical*; *n* = 36; *age* = 20.89 ± 0.92) were clustered under the PE subclinical group [37]. All these individuals were sourced from Avon Longitudinal Study of Parents and Children (ALSPAC) birth cohort [38]. Randomly selected controls (*n* = 269; *age* = 22.30 ± 1.46), from the same cohort who had undergone the same assessments but who were rated as not having had PE experiences, were also scanned.

MRI data from first-degree relatives of schizophrenia (SCZ-relatives; *n* = 96; *age* = 43.75 ± 15.25) and schizoaffective disorder (SAD-relatives; *n* = 64; *age* = 40.00 ± 16.20), clustered under SCZ and SAD-relatives group; were obtained from B-SNIP. The HC group consisted of the same individuals as the chronic group.

All datasets and protocols have been approved by the corresponding ethical committees, and written informed consent was received prior to participation. Additional inclusion and exclusion criteria can be found in *Supplementary Subjects and Methods*. See Supplementary Table 1 for demographic details, and Supplementary Fig. 1 for age distributions for each diagnosis and dataset.

### MRI acquisition, parcellation, and volume extraction

High-resolution brain MRI scans were obtained on different MRI scanners (1.5 – 3T) and acquisition protocols (see *Supplementary Subjects and Methods* for details). T_1_-weighted images were acquired with sequences tailored to the respective scanner specifications, and were processed using FreeSurfer (http://surfer.nmr.mgh.harvard.edu) applying the recon-all pipeline to enhance gray-white matter boundary delineation. In cases where raw T_2_-weighted were also available, T_1_-T_2_ recon-all pipeline was applied.

Cortical brain parcellation was performed using Desikan-Killiany (DK) atlas, and volumetric measurements were derived for each region-of-interest. Quality control procedures were implemented to ensure the accuracy and reliability of the derived cerebral volumes. Thirteen HC subjects exhibiting significant artifacts or processing errors were excluded from further analysis.

### Centile and effect sizes estimation and analysis

To benchmark regional volumes of each psychosis-related diagnosis and group against normative trajectories, we used a generalized additive model for location, scale, and shape (GAMLSS) [40]. This model, available at https://github.com/brainchart/Lifespan, estimated cross-sectional normative age-related trends from 100 different studies (around 120,000 participants; see Supplementary Information in [20] for details of the normative cohort, pages 120-151). Therefore, age-normed and sex-stratified measures of brain structure atypicalities across the lifespan, known as centiles (Fig. 1A), could be derived for each psychosis-related and HC individual in our cohorts (Fig. 1B; see Supplementary Subjects and Methods for details). For example, a subject at the 20^th^ centile would have a volume that is “approximately” lower than 80% of individuals of the same age and sex after adjusting for scanning site offset. Since the models provided by Bethlehem *et al*. [20] were only available for brain volume averaged across hemispheres, only 34 DK regional centiles (instead of 68) could be derived for each individual.

**Fig. 1.**
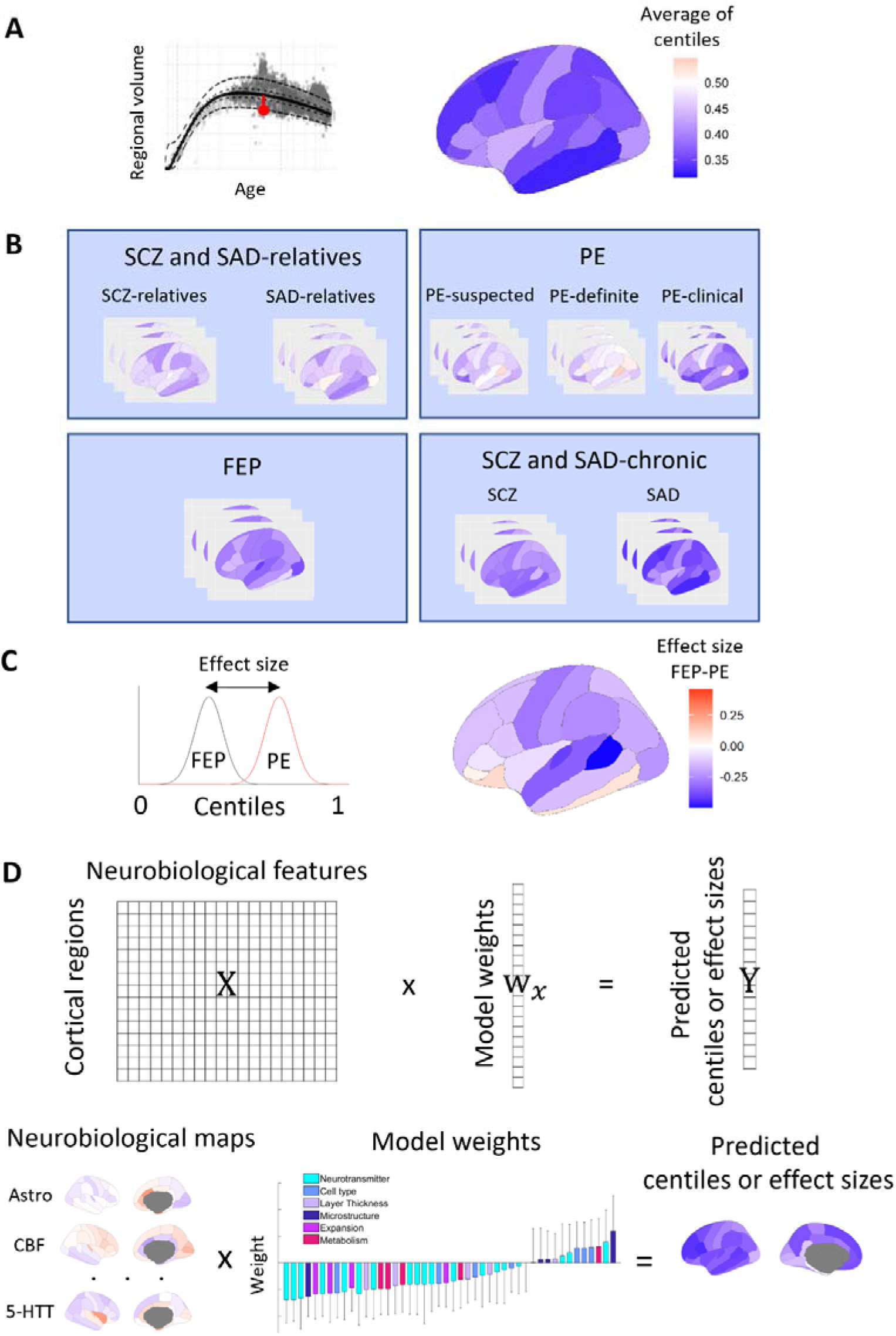
Analysis pipeline. Centiles and effect sizes were computed from regional brain volumes, which were then predicted based on combinations of neurobiological features. (**A**) Deviation of regional volume from the median volume of neurotypical population for a single patient (red dot). Resulting ranked deviations, known as centiles, were computed for individuals with the same diagnosis for each brain region defined in the Desikan-Killiany atlas. Centiles range from 0 to 1, with values below 0.5 indicating that an individual has a lower GM volume than the expected normative values for their age and sex. (**B**) MRI data analyzed in the present study included eight different psychosis-related diagnoses clustered into four groups according to their clinical profile. (**C**) Effect sizes were computed as the Cohen’s d between regional centiles for each pair of groups. (**D**) Associations between neurobiological maps and empirical centiles (or their effect sizes) were conducted using PCA-CCA modeling which resulted in a set of predicted centiles (or effect sizes) derived from linear combinations of neurobiological features.

Regional centiles of each diagnosis, calculated as the mean across individuals within the same diagnosis, were compared with HC using the Wilcoxon rank-sum test. Additionally, Benjamini-Hochberg false discovery rate (FDR) correction across brain regions was applied to account for multiple comparisons. Centile distributions of each diagnosis were averaged again with those similar diagnoses according to their clinical profile to constitute each of the four psychosis-related groups considered here (SCZ and SAD-relatives, PE, FEP, and SCZ and SAD-chronic). Global centiles (i.e., mean centile across regions) and regional centiles of each group were also compared with HC using the Wilcoxon rank-sum test. Additionally, differences between psychosis-related groups were assessed by calculating for each pair of groups: (1) the effect size (Cohen’s d) of centiles (Fig. 1C); and (2) the Sum of Squared Differences (SSD) between them. To statistically assess these differences, a permutation test (FDR-corrected) was applied for each pair of group comparisons by randomly shuffling group assignment to create a null distribution (10,000 permutations). Additionally, centiles are highly robust to variations in image quality (see Supplemental Material of [20]). Nevertheless, sensitivity analyses using the Euler index as a proxy for image quality were conducted to evaluate the stability of centile estimations.

### Neurobiological cortical maps

We expanded the methodology proposed by Hansen *et al.* [41] to explore potential associations between cortical volume centile profiles and the spatial maps of 46 molecular and micro-architectural attributes (collectively referred here as neurobiological features) collected across multiple studies. These maps were obtained in surface or volumetric spaces using *neuromaps* toolbox, available at https://github.com/netneurolab/neuromaps, and were parcellated according to DK atlas. In cases where multiple maps were available for a single feature, a weighted average was taken. Neurobiological maps were classified under six different types of neurobiological features (see Supplementary Table 2 for a complete list of neurobiological features) [42]: neurotransmitter (19 features), cell type (7), layer thickness (6), microstructure (5), cortical expansion (4), and metabolism (5).

### Principal Component Analysis - Canonical Correlation Analysis (PCA-CCA)

A combined Principal Component Analysis (PCA) and Canonical Correlation Analysis (CCA) approach was employed to relate neurobiological maps to regional centiles. CCA is a powerful multivariate method for capturing associations between two modalities of data (e.g., brain and behavior) [43]. However, when the sample size (34 cortical regions) is similar to or smaller than the number of variables (46 neurobiological features), standard multivariate models may overfit. To address this problem, a prior dimensionality reduction with PCA was performed. For this purpose, PCA-CCA was executed for different explained variances (60, 70, 80 and 90%). To assess the statistical significance of the models, a spatial autocorrelation-preserving permutation test, termed ‘spin test’, was used for each of the four variances considered (see *Supplementary Subjects and Methods* for details). To minimize the risk of overfitting, the model selected for each PCA-CCA analysis was the one with the lowest explained variance that was significant according to the spin test (FDR-corrected; see Supplementary Data). For non-significant models, the model with lowest explained variance (60%) was selected for illustrative purposes.

For each of the psychotic group analyses, the PCA-CCA model identified a set of weights (w_x_) that the resulting linear combination (weighted sum) of the neurobiological maps constituted a set of predicted centiles that are, by construction, correlated with the empirical centiles (Fig. 1D). To assess the significance of the model weights, the autocorrelation-preserving spin test was employed. To estimate the standard errors, we created 1,000 bootstrap samples by sampling with replacement the observations of the molecular maps. Finally, to ascertain the extent to which each neurobiological feature contributed to the predicted centiles, a set of loadings was computed as the Pearson correlation between the neurobiological map and the dimensionally-reduced predicted centiles.

### Neurobiological similarity and structural co-vulnerability to psychosis matrices

Neurobiological features were correlated across regions to obtain a region-by-region matrix of “neurobiological similarity”. Simultaneously, a 34-by-4 matrix was constructed to represent structural disorder abnormality based on the 34 regional effect sizes of the centiles for each of the groups (SCZ and SAD-relatives, PE, FEP, and SCZ and SAD-chronic) with respect to the HC group. This matrix was further correlated to evaluate the extent to which each pair of regions exhibited similar effects across different groups, resulting in a 34-by-34 matrix referred to as “structural co-vulnerability to psychosis”. Subsequently, both the “neurobiological similarity” and “structural co-vulnerability to psychosis” matrices were correlated to identify associations (spin-tested) between regions with similar neurobiological attributes and overlapping structural vulnerability profiles (centiles). Thus, a pair of neurobiological features co-localized in the same region would be associated with a pair of structurally altered regions that tend to vulnerate to psychosis in a similar way. The same procedure was followed to represent structural disorder abnormality based on the regional effect sizes of the centiles for each diagnosis (rather than for each group).

## Results

The normative approach assigned ranked centiles to regional brain volumes of each individual, identifying deviations from the expected normative trajectories while accounting for age, sex, and site-effects (see Supplementary Figs. 2 and 3 for sex and site-effects, respectively). Thus, after averaging brain volumes across hemispheres for each of the 34 Desikan-Killiany regions, centiles were computed for 38,696 HC and 1,256 individuals classified under eight different psychosis-related diagnoses, which were further clustered into four groups according to their clinical profile: *SCZ and SAD-relatives*, *PE*, *FEP*, and *SCZ and SAD-chronic*.

### Reduction of regional centiles in psychosis-related conditions

We initially computed the mean regional centile distribution (i.e., centile maps) for eight psychosis-related diagnoses and the four groups in which they were clustered (Fig. 2). We found significantly decreased centile scores in global GM volume across all groups (Wilcoxon rank-sum test FDR-corrected; relatives *P* = 0.0021; PE *P* = 0.0124; FEP *P* < 10^-7^; chronic *P* < 10^-28^). The effect of global centiles, as well as the quality image (Euler index), did not alter these findings (see Supplementary Figs. 4 and 5, respectively). Compared to HC, SCZ-relatives revealed several significantly reduced regions in association cortices. Although SAD-relatives did not show significant differences in centiles compared with HC, the group that comprised both SCZ and SAD-relatives altogether exhibited a greater number of broadly distributed significant reductions. Individuals with ‘suspected’ and ‘definite’ PE classifications showed no significant differences in centiles compared to HC, whereas the PE-clinical group exhibited 11 regions significantly reduced. The FEP group, however, exhibited significant differences in all regions compared to HC, except for the inferior temporal and the temporal pole. Lastly, SCZ showed significantly reduced centiles for all regions, while SAD exhibited significant decreases in the majority of them. The within-condition centile variability was similar for all diagnoses and groups (see regional standard deviations in Supplementary Fig. 6). The extremely deviant regions for each group, calculated as the relative frequency of individuals with a regional centile < 0.05, were similar to the decrease in mean centiles described in Figure 2 for PE, FEP, and SCZ and SAD-chronic groups (Supplementary Fig. 7).

**Fig. 2.**
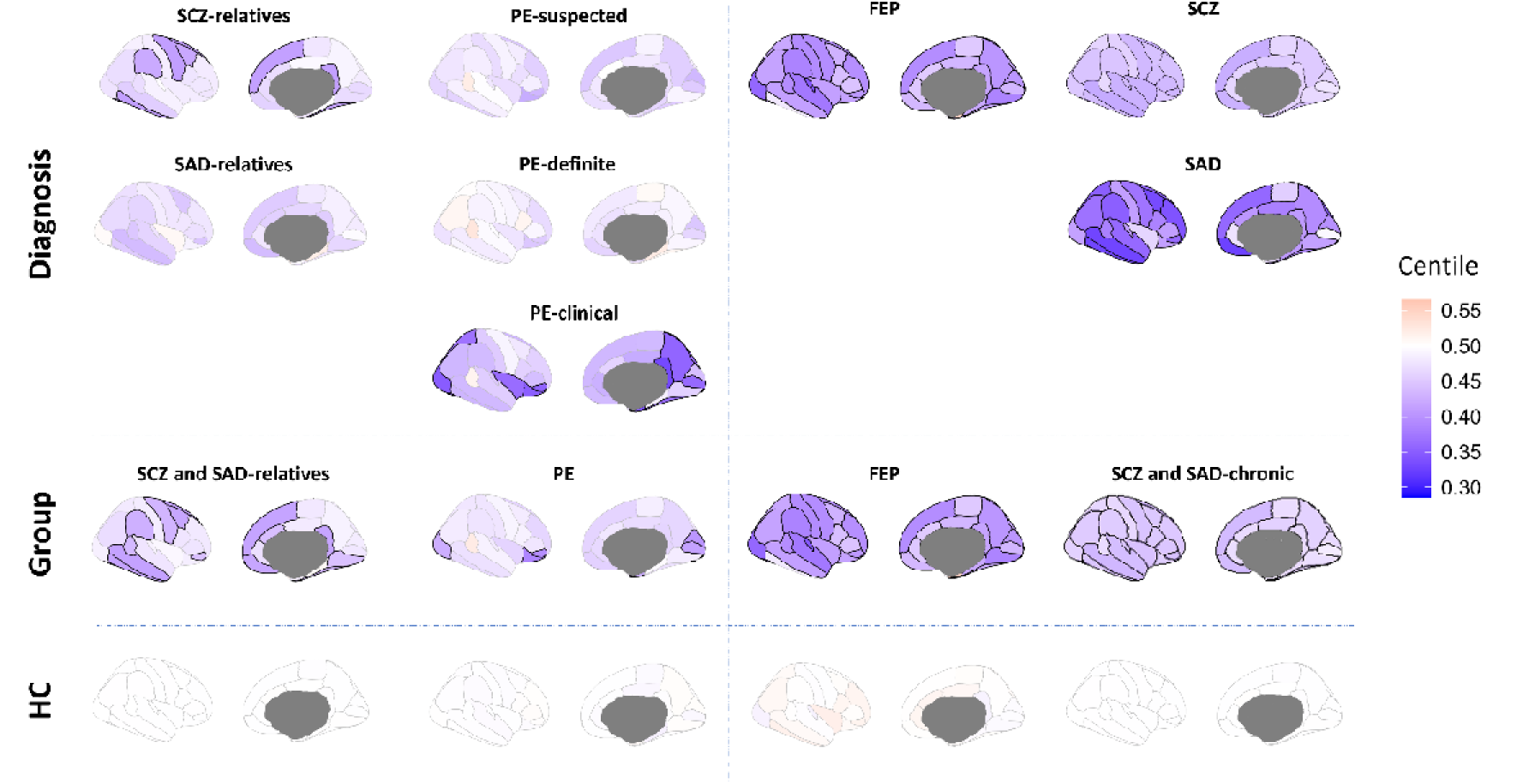
Regional brain volume centiles. Regional MRI brain volumes were converted into centiles and subsequently averaged across individuals to generate a mean centile map for each diagnosis and group. The highlighted regions show those regional centiles that exhibit significant differences from HC after FDR correction (Wilcoxon rank-sum test, *P* < 0.05).

### Differential effect sizes of regional centiles between psychosis-related groups

To determine how centiles vary between psychosis-related groups, we compared the effect size (Cohen’s d) between the regional centiles of each pair of psychosis-related groups (Fig. 3). Additionally, we assessed the similarities between centiles of the compared groups by computing the Sum of Squared Differences (SSD) across regions.

**Fig. 3.**
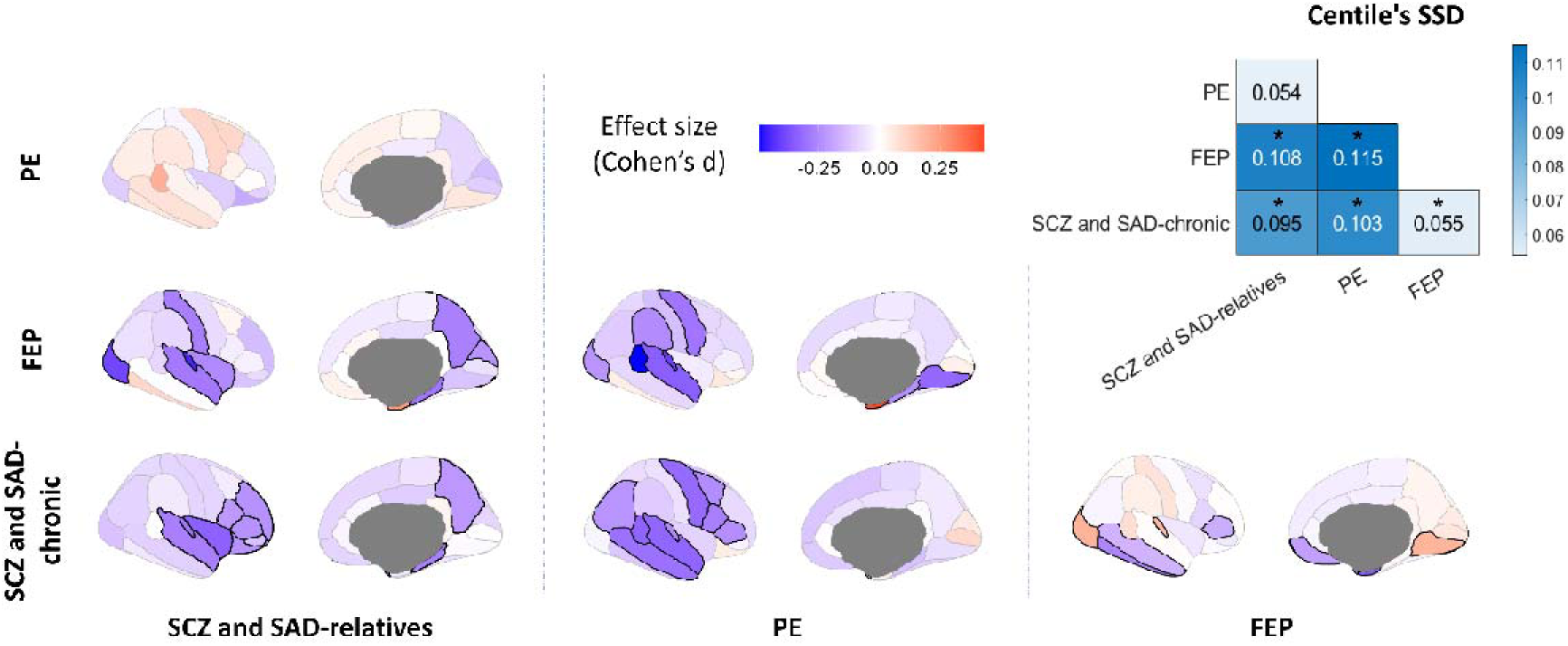
Effect sizes of centiles between groups. Cohen’s d was computed between regional centiles of each pair of groups to map the effect sizes of centiles between conditions. The highlighted regions show those regional effect sizes that exhibit significant differences between groups after FDR correction (*P*_perm_ < 0.05). Top-right panel represents the sum of regional centile squared differences (SSD) between groups. Asterisks (*) indicate significant differences in SSD between groups (FDR-corrected *P*_perm_ < 0.05).

All pairs of groups showed significant differences in centile distributions (Fig. 3 top-right corner; all *P*_perm_ < 0.005), except for SCZ and SAD-relatives versus PE (*SSD* = 0.054, *P*_perm_ = 0.166). These generalized differences in centiles were also supported by the low regional Pearson correlation between groups (Supplementary Fig. 8; all *P*_perm_ > 0.05). At the regional level, the chronic group exhibited decreased centile values in frontal and temporal lobes compared to the relatives, PE, and FEP. However, in comparison to FEP, the chronic group also showed an increase in the occipital lobe and the transverse temporal region. Lastly, FEP group demonstrated increased centile values in entorhinal, and a decrease in frontal, temporal, and occipital lobe regions compared to the relatives and PE groups.

### Mapping neurobiological maps to centiles

A combined PCA-CCA approach was used as a multivariate method to capture associations between neurobiological maps (*X*) and regional centiles (*Y*; Fig. 1D) [43]. Predicted centiles resembled empirical centiles for statistically significant models (Fig. 4A; all groups except PE; FDR-corrected *P*_spin_ < 0.05; see Supplementary Data). The two clinical groups that showed the lowest regional centiles, FEP, and SCZ and SAD-chronic, also revealed the strongest correlation between predicted and empirical centiles (Fig. 4B; *r* = 0.63 and *r* = 0.68, respectively). All significant models exhibited significant loadings (Fig. 4C; *P*_spin_ < 0.05). Specifically, all loadings were negative for the relatives, indicating that the presence of these neurobiological features is highly co-localized with regions that exhibit low centiles. Groups with the lowest centiles, FEP and chronic, showed a greater number of significant loadings, most of which were found to be negative. Synapse density and 5-HT_2A_ demonstrated a large negative contribution for all significant groups. Cortical expansion (Evolutionary exp., Scaling NIH, and Scaling PNC) and neurotransmitters (5-HT_2A_, α_4_β_2_, mGluR_5_) provided a higher number of negative loadings for the relatives and the chronic group, indicating a higher presence of these features in regions with lower centiles. However, metabolism (CBF, CMRO_2_, CMRGlu) and microstructure (Gene PC1, Myelin (T_1_-w/T_2_-w), and Synapse density) predominated negatively in FEP group. Layer thickness loadings (Layers I, II, V, VI) made a significant negative contribution in the chronic group. Conversely, positive loadings indicated a high presence of these neurobiological features in regions closer to neurotypical (higher) centiles, or equivalently, a low presence in affected regions (lower centiles). Cell type loadings were both positive (low presence of Micro, OPC, Astro at low centile regions) and negative (high presence of Neuro-Ex, Neuro-In at low centile regions) in FEP. In contrast, 5-HT_6_, 5-HTT, D_1_, D_2_, DAT, H_3_, NMDA, VAChT, Endo, Oligo, Layer III, and Neurotransmitter PC1 did not exhibit significant associations with differences in centile for any of the psychosis-related groups. These loading distributions were similar to those of the weights of the models (Supplementary Fig. 9). The effect of parcel size (DK atlas) did not alter these findings (see Supplementary Fig. 10 for a subdivision of the DK atlas into Schaefer’s 400 parcellation). As a sensitivity test, we performed a Partial Least Square (PLS) analysis to assess whether the loading estimations were highly influenced by the PCA-CCA method used here. A high correlation (r > 0.92) between loadings derived from both methods was found (Supplementary Fig. 11). Leave-one-study-out cross-validation performed both within the chronic group and SCZ diagnosis indicated that centiles and loadings were not primarily influenced by individual studies (Supplementary Figs. 12 and 13, respectively). Additionally, empirical and predicted centiles, and associated loadings of each individual diagnosis demonstrated a high consistency across diagnoses classified under the same group (Supplementary Figs. 14-16). The associations between the neurobiological maps (*X*) and the effect sizes of centiles between each pair of groups (*Y*), captured by the loadings, showed significant differences in neurobiology between all conditions except those involving relatives (see Supplementary Fig. 17). These loading distributions were similar to those of the weights of the models (Supplementary Fig. 18).

**Fig. 4.**
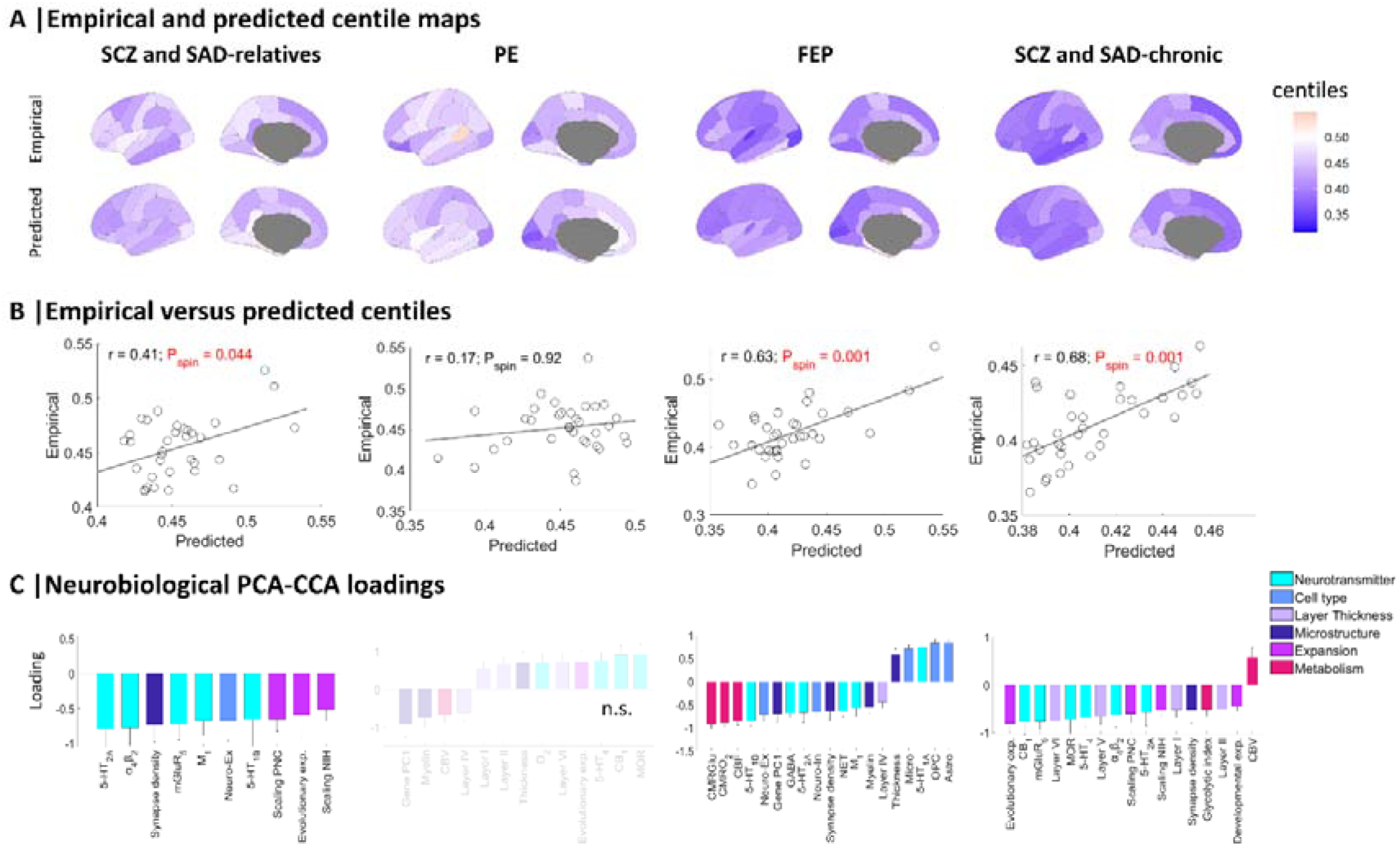
Empirical and predicted centiles, and associated loadings from PCA-CCA models. (**A**) Maps of empirical MRI-derived centiles (top) and predicted PCA-CCA-derived centiles from neurobiological features (bottom). (**B**) Correlation between empirical and predicted regional centiles. (**C**) PCA-CCA significant loadings associated to each neurobiological map (*P*_spin_ < 0.05). Non-significant models are denoted as n.s (FDR-corrected *P*_spin_ > 0.05). Error bars represent the standard deviation.

### Comparing neurobiological loadings across psychosis-related groups

We next explored the consistency of loadings described in Fig. 4 by stacking them across groups to reveal potential overlapping features across different conditions (Fig. 5A). The greatest negative loadings indicated a high presence of these features in regions where centiles are consistently low across the four groups. These features included neurotransmitters (from high to low model contribution: 5-HT_1B_, 5-HT_2A_, α_4_β_2_, NET, GABA), cell types (Neuro-Ex), microstructure (Synapse density), and metabolism (CMRGlu, CBF, and CMRO_2_). On the other hand, the greatest positive loadings were consistent across all groups except chronic. These features included neurotransmitters (5-HT_1A_, 5-HTT, D_2_, DAT), cell types (Astro, OPC, Micro), and layer thickness (Layer III). In contrast to the less severe cases, the chronic group mainly exhibited negative loadings, indicating a generalized presence of features in regions with decreased centiles. These loading distributions were similar to those of the weights of the models (Supplementary Fig. 19).

**Fig. 5.**
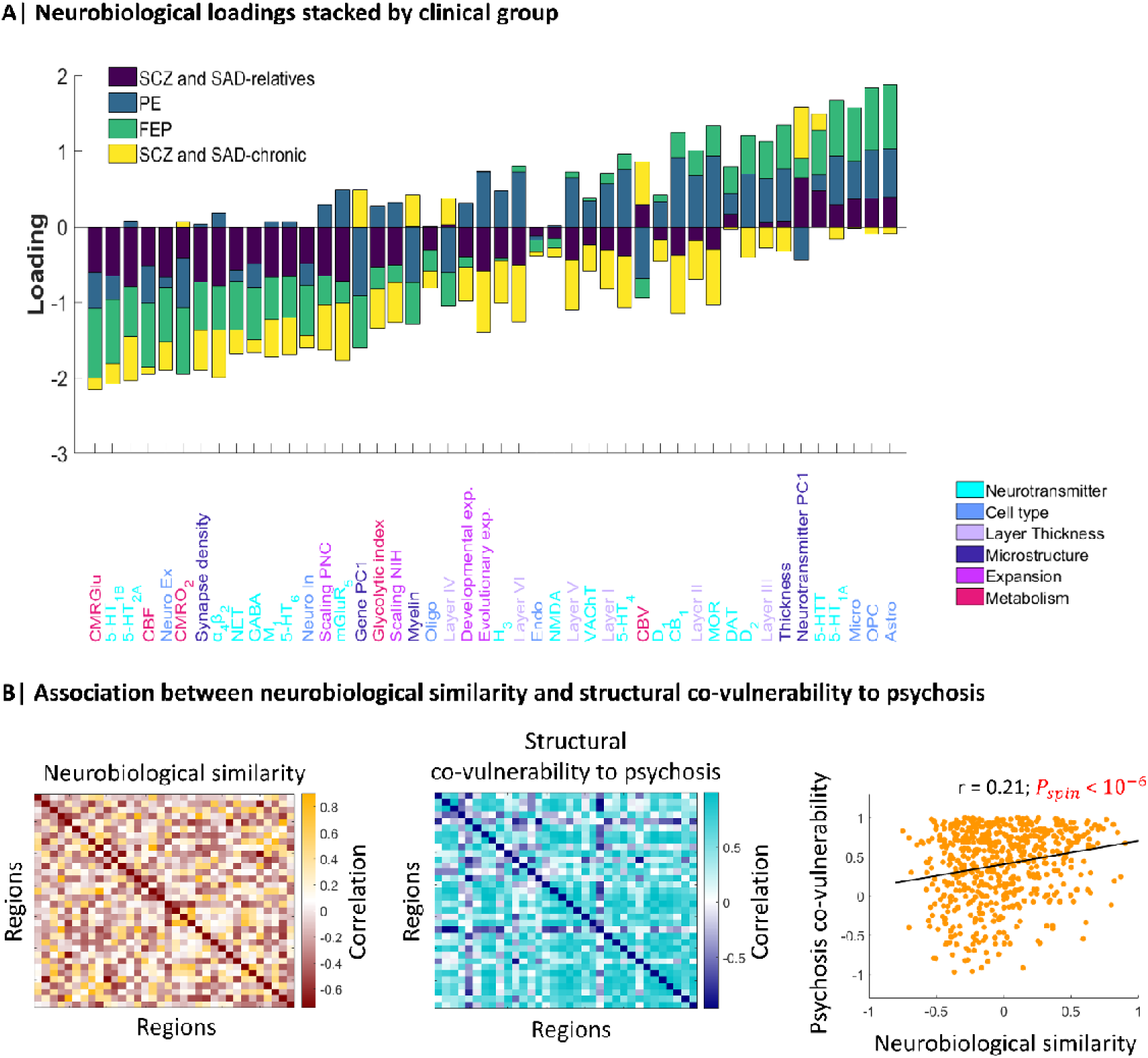
Shared neurobiological features among psychosis-related groups. (**A**) Stacked neurobiological loadings of each group, regardless of their significance, were ranked from the most negative to the most positive average contribution. (**B**) Neurobiological similarity matrix obtained by correlating the regional patterns of neurobiological features in HC (left). Structural co-vulnerability to psychosis matrix constructed by correlating the regional patterns of the effect sizes of centiles across psychosis-related groups (middle). Association between neurobiological similarity and structural co-vulnerability to psychosis (right).

This shared neurobiological spatial distribution across different psychosis-related groups prompted us to investigate whether regions with similar neurobiological attributes also tend to exhibit overlapping structural vulnerability profiles (centiles). Thus, we created a neurobiological similarity matrix and a structural co-vulnerability to psychosis matrix, which showed a common inter-regional centile reduction across the psychosis spectrum (Fig. 5B; median *r* = 0.52 ± 0.32). The correlation between both matrices revealed a significant association between the neurobiological similarities and the structural co-vulnerabilities to psychosis (Pearson’s *r* = 0.21, *P*_spin_ < 10^-6^). A significant correlation was also found between the neurobiological similarities and the effect sizes of centiles when considering individual diagnoses instead of groups (Supplementary Fig. 20). This association was also found when considering only the most contributing features (see Supplementary Figs. 21 and 22).

## Discussion

In the present report, we employed a centile-based method to identify, under the neurodevelopmental hypothesis of psychosis, cortical volume deviations below the expected maturational trajectory for different groups of psychosis spectrum (SCZ and SAD-relatives, PE, FEP, and SCZ and SAD-chronic). The predictions of PCA-CCA models captured associations between neurobiological maps and the reduced centiles. This resulted in a set of loadings that reflected how structural differences were co-localized with a set of neurobiological features, providing additional support for the regional vulnerability hypothesis. Accordingly, regions with similar neurobiological attributes also tended to exhibit overlapping profiles of structural vulnerability to psychosis.

Age- and sex-related neurodevelopmental events play a major role in psychotic conditions [15]. Using a centile method that removes age-, sex-, and site-effects [40], we were able to detect brain volume deviations from the expected trajectories. A key benefit of this method, compared to other normative models used in schizophrenia [44, 45], lies in its extensive dataset (more than 120,000 scans) that spans the entire lifespan, from 17 post-conception weeks to 100 years. Thus, our centile-based study provides a standardized and interpretable measure of regional brain volume atypicalities for unveiling patterns of neuroanatomical differences across psychiatric disorders that emerge during development and aging. Here, centiles revealed a significant volume decrease in all psychosis-related groups, with a greater impact on the most severe conditions, FEP and chronic. These results support a continuum view of psychosis psychopathology, as the number of significantly affected regions increased with the severity of the disorder. In particular, the clinically diagnosed individuals (FEP, SCZ, and SAD) exhibited overlapping GM reductions in frontotemporal and anterior cingulate cortices, which aligns with the reported results in several studies [9, 46, 47]. Interestingly, pars orbitalis, a region that is related to mechanisms that lead to volumetric decrease in other cortical regions in SCZ [48], displayed a significant volume reduction in all groups. Decreased centiles shown by relatives compared with HC supports the hypothesis that cortical regions such as cingulate, temporal, and frontal regions may be preferentially affected by the latent genetic liability to SCZ in these asymptomatic individuals [11, 49]. On the other hand, the PE group did not show any significant reduction in centiles (only PE-clinical individuals), consistent with previous voxel-based morphometry analyses of the same individuals [50]. This may be attributed to the population diversity (suspected, definite, and clinical PEs, which exhibited heterogeneous centile distributions) and the subclinical nature of the symptoms (only some patients had recurrent PEs) [51]. We also did not find significant centile differences between the two subclinical groups considered: relatives and PE. All other pairs of groups showed significant differences in centile distribution. The decreased centile values in frontal and temporal lobes of the chronic group, compared to the relatives and PE, are consistent with the stage and severity of the disorders as previously reported [47]. Compared to FEP, the chronic group showed a decrease in centiles in temporal lobe, and an increase in the occipital lobe. The decrease aligns with the progressive reduction of frontotemporal regions described in individuals with long-term SCZ [52]. The GM increase in the occipital lobe has also been reported in SCZ patients compared to relatives [53] and to healthy controls [54]. This has been interpreted as a result of brain plasticity attempting to compensate for reduced connectivity [54]. Lastly, the decrease in centiles in the frontal, temporal, and occipital lobes of FEP group compared to relatives and PE groups, supports the relevance of these regions in GM reduction in this early stage of psychosis-related diseases [8, 55]. Collectively, these findings highlight the presence of reduced centile patterns within each psychosis spectrum disorder, emphasizing the specific and prominent role that abnormal brain maturation plays in the severity of psychosis.

The diagnosis and treatment of these conditions has often presented a challenge due to the heterogeneous nature of these mental disorders, their shared symptomatology, and the limited understanding of the underlying neurobiological mechanisms [3]. Reductions in GM volume have been associated with several neurobiological features, including variants of the serotonin transporter gene [56], reductions in synaptic density [57] and myelination [58], increases in glucose metabolism [59], as well as astrogliosis and pro-inflammatory cytokines produced by neurons, astrocytes, microglia, and oligodendrocytes [60]. Here, we employed a PCA-CCA multivariate method that assessed differential regional GM vulnerabilities across psychosis-related diagnoses by associating cortical volumes with neurobiological features derived from humans. In this line, recent research has co-localized structural brain development with the underlying neurobiology [61]. Furthermore, the spatial distribution of neurotransmitter receptors and transporters [41], and the connectional hierarchy [62] have been associated with cognitive processes and disease vulnerability. Additionally, a study has suggested that regions that are structurally most vulnerable to disease may also be the most susceptible to rebalance their functional organization through appropriate pharmacological interventions [63].

The large negative contribution of synapse density and 5-HT_2A_ serotonin receptor in all psychosis-related groups indicated a high presence of these features in neurotypical regions that exhibit reduced centiles due to psychosis, supporting the regional vulnerability hypothesis. For example, we found psychosis-related centile reductions in areas with higher synapse density, which is consistent with the loss of synaptic density detected in SCZ patients [57]. An abnormal expression pattern of 5-HT_2A_ has been suggested to predispose an individual to the development of psychosis [64] as well as being implicated in the pathogenesis of suicidal behavior through genetic associations in patients with SCZ [65]. In this line, an inverse agonist of 5-HT_2A_ has been discovered to reverse psychosis-like behaviors in a rodent model of Alzheimer’s [64] and Parkinson’s disease [66]. Additional neurotransmitters that made a negative contribution to the relatives and chronic models included α_4_β_2_ nicotinic acetylcholine receptor. Studies in rodents have shown that α_4_β_2_ agonists enhance sensory gating, an information processing function that is deficient in SCZ [67]. Pharmacological research in animals has also indicated that α_4_β_2_ is involved in multiple cognitive domains impaired in SCZ, including processing speed, visual learning and memory, and social cognition [68]. Pre-clinical evidence has suggested that agonists of the nicotinic α_4_β_2_ subtype could be beneficial in improving cognitive function in individuals with SCZ. We also found consistencies regarding microstructural features beyond synapse density in the FEP group, where negative loadings predominated. According to the literature, changes in cortical myelination and cortical thickness are co-localized with the expression of genes associated with SCZ [22, 70]. The remarkable negative contribution of brain metabolism in FEP may represent a complementary vulnerability mechanism to SCZ. Brain-metabolic features have demonstrated to explain a substantial amount of the variance associated with regional cortical thickness trajectories during childhood and adolescence [61]. Specifically, altered CBF has been found in people at risk for psychosis, in FEP and in SCZ [71]. The negative loading contribution of cortical expansion features in relatives and the chronic group suggests that allometric scaling directly parallels the incidence of neurodevelopmental impairments, as demonstrated in preterm infants [72]. Furthermore, major contributor genes to the rapid evolutionary expansion of the human brain are also significant contributors to SCZ [73]. Layer thickness, a feature related to neuronal density [74], also had a significant negative contribution in the chronic group. Moreover, we reported that several cell type loadings contributed positively in FEP. Thus, a high presence of these cell types prevails in regions that closely align with neurotypical centiles, suggesting that their absence might be a contributing factor to vulnerability to the disease. This finding supports the increased complement-mediated microglial pruning and the enhanced phagocytic action of microglial cells produced by astrocytes that has been reported at the onset of SCZ [75].

Beyond the contribution of neurobiological features found across psychotic conditions, we identified a consistent pattern of overlapping negative loadings across all groups, including neurotransmitters, synapse density, and metabolism. Positive loadings included neurotransmitters, cell types, and Layer III for all groups except chronic. This overlapping pattern across all groups indicates a high consistency in the features that may be partially responsible for vulnerability to psychosis, supporting the hypothesis of a continuum spectrum for the disorder. The predominance of negative loadings in the chronic group is interpreted as a manifestation of a higher neurobiological vulnerability to brain alterations, which is associated with the severity of this stage. This shared neurobiological vulnerability across different psychosis-related groups led us to investigate the relationship between regions with similar neurobiological attributes and overlapping structural vulnerability profiles (centiles). The significant correlation between the neurobiological similarities and the structural co-vulnerabilities to psychosis revealed that pairs of regions sharing neurobiological profiles tend to exhibit comparable vulnerabilities across psychotic conditions. In this line, Luppi *et al*. [63] recently found that inter-regional neurotransmitter similarity was associated with pharmacological susceptibility which, in turn, correlated with a vulnerability pattern to neurological, neurodevelopmental, and psychiatric conditions.

The present findings must be interpreted with several considerations. First, psychotic disorders were grouped with varying sample sizes and diagnoses, resulting in heterogeneous groups. Nevertheless, it has been suggested that SCZ and SAD patients exhibit overlapping areas of GM reduction [13], and these disorders are even considered neuropsychologically indistinguishable [12]. Additionally, we replicated our findings by considering individual diagnoses instead of groups (Supplementary Figs. 14-16). Second, while abnormalities in relatives, PE, and FEP individuals are not influenced by medication, antipsychotic drugs may impact cortical volume in chronic individuals [75]. Third, since computing the normative trajectories requires hundreds of thousands of CPU hours, the models provided by Bethlehem *et al*. [20] were only available for cortical brain volume averaged across hemispheres (not subcortical volume, cortical thickness, or surface area). Fourth, although in this limited sample size scenario (34 regions) the CCA multivariate approach has a risk of over-fitting [76], we first applied PCA before CCA to prevent this issue, and then performed a PLS to confirm the results. Fifth, neurobiological data used here were derived from normative non-psychotic individuals. Future work with a neurobiological atlas of individuals with schizophrenia could provide new insights into the molecular alterations causally associated with abnormal brain alterations. Sixth, model weights were formulated to maximize the association between neurobiological features and centiles. However, we opted to report loadings instead, as they depict the extent to which each feature contributed to the model. Nevertheless, loadings and weights exhibited high resemblance (compare Fig. 4C and Supplementary Fig. 9; Supplementary Fig. 17 and Supplementary Fig. 18; and Fig. 5A and Supplementary Fig. 19).

In summary, we identified group-specific volume deviations below the expected trajectory for different psychosis-related conditions based on centiles. We revealed an overlapping spatial distribution of the neurobiological features, which were highly co-localized with the abnormal GM trajectories. Altogether, these findings contribute to our understanding of the vulnerability factors that may underlie atypical brain maturation in different conditions and stages of psychosis, which could help define subtypes for future imaging-first molecular phenotyping.

## Acknowledgements

NGS and RRG are funded by the EMERGIA Junta de Andalucía program (EMERGIA20_00139). RRG is also supported by the *Plan Propio* of the University of Seville, the *Plan de Generación de Conocimiento* (PID2021-122853OA-I00) and the *Plan de Consolidación* (CNS2023-143647) from the Agencia Estatal de Investigación.

## Data availability

Volumetric MRI images from the ALSPAC dataset are available at https://www.bristol.ac.uk/alspac/researchers/access/. PAFIP data are available from the corresponding author on request. The ABCD dataset is available at https://nda.nih.gov/abcd/. ASRB data is supported by Neuroscience Research Australia (NeuRA), available at https://neura.edu.au/resources-tools/asrb. The B-SNIP dataset is available at https://nda.nih.gov/edit_collection.html?id=2274. The LA5c dataset was obtained from the OpenfMRI database (https://legacy.openfmri.org/dataset/ds000030/). MCIC data is available at https://coins.trendscenter.org/, and UKB dataset at https://www.ukbiobank.ac.uk/. The Desikan–Killiany parcellation atlas was obtained from netneurotools (https://github.com/netneurolab/netneurotools). All code used to perform the analyses can be found at https://github.com/RafaelRomeroGarcia/NeurobiologyCentilesPsychosis. The code used for the PCA-CCA analyses is available at https://github.com/RafaelRomeroGarcia/cca_pls_toolkit.

## Supplementary Information

Supplementary information is available at MP’s website.

**This PDF file includes:**

Supplementary Subjects and Methods

Supplementary Figs. 1 to 22

Supplementary Tables 1 and 2

**Other Supplementary Materials for this manuscript include the following:**

Supplementary Data (.xlsx)

