## Supplemental Material for "Molecular and micro-architectural mapping of gray matter alterations in psychosis"

**This PDF file includes:**

Supplemental Subjects and Methods	2

**Datasets description**	2

ABCD – Adolescent Brain and Cognitive Development	2

ALSPAC – Avon Longitudinal Study of Parents and Children	3

ASRB – Australian Schizophrenia Research Bank	4

B-SNIP – Bipolar & Schizophrenia Consortium for Parsing Intermediate Phenotypes	5

LA5c – UCLA Consortium for Neuropsychiatric Phenomics LA5c Study	6

MCIC – Mental Illness and Neuroscience Discovery (MIND) Institute Clinical Imaging Consortium	7

PAFIP – Programa de Atención a las Fases Iniciales de Psicosis	9

UKB - UK Biobank	10

**Spin test**	11

Supplemental References	12

Supplemental Figures	15

Supplemental Tables	36

**Other Supplementary Information for this manuscript includes:**

Supplementary Data (separate .xlsx file)

### Supplemental Subjects and Methods

#### **Datasets description**

##### ABCD – Adolescent Brain and Cognitive Development

The ABCD study (NDA ID:2668) is a landmark, longitudinal study of brain development and child health [1]. A baseline cohort of 11,500 nine- and ten-year-old children (and their parents/guardians) are being recruited and will be followed for ten years with annual lab-based assessments including biennial MRI [2]. The study evaluates brain maturation within the context of social, emotional, and cognitive development, as well as a variety of health and environmental outcomes. ABCD Study employed probability sampling of U.S. schools within the 21 catchment areas as the primary method for contacting and recruiting eligible children and their parents. Sampling of schools and consenting students/parents has also been used to recruit cohorts for a number of major national studies. Individuals were included in the reference model as healthy controls based on the parental response to the ABCD screening and risk questionnaire (<https://nda.nih.gov/data_structure.html?short_name=abcd_screen01>) indicating the individual had never been diagnosed with a mental health disorder.

The ABCD Study builds upon existing state-of-the-art imaging protocols from the Pediatric Imaging, Neurocognition, and Genetics (PING) study [3], and the Human Connectome Project (HCP) [4] for the collection of multimodal data: T_1_-weighted and T_2_-weighted structural MRI (sMRI), diffusion MRI (dMRI), and functional MRI (fMRI), including both resting-state fMRI (rs-fMRI) and task-fMRI [5].

MRI data were acquired using 3T MRI scanners (*Siemens Prisma and Prisma Fit*, *GE MR 750*, and *Philips Achieva dStream and Ingenia*) [5]. The ABCD imaging protocol was harmonized across data collection sites for three 3T scanning systems, all of which used standard adult-size multi-channel head coils and multiband echo planar imaging acquisitions [6]. High-resolution structural imaging sequences, such as T_1_-weighted (1.0 mm isotropic voxels) and T_2_-weighted axial fast-spoiled gradient echo sequence (repetition time [TR] = 2400-2500 ms, echo time [TE] = 2-2.9 ms, matrix size = 256×256, field-of-view [FOV] = 256×240-256 mm^2^, flip angle = 8°, inversion delay = 1060 ms, and 176-225 sections), were employed to capture detailed information of the brain anatomy. Images were corrected for distortions and head motion. FreeSurfer version 6.0.1 software (<http://surfer.nmr.mgh.harvard.edu>) [7] was used to perform the cortical reconstruction and volumetric segmentation, and cortical or subcortical regions were parcellated and labeled with atlas classification.

##### ALSPAC – Avon Longitudinal Study of Parents and Children

ALSPAC (<https://www.bristol.ac.uk/alspac/researchers/access/>) is a pregnancy and birth cohort established to identify the factors influencing child health and developmental outcomes [8]. The study is composed by pregnant women from Avon, South West of England, with an expected date of delivery between 1 April 1991 and 31 December 1992, who had completed the questionnaire scheduled for the third trimester of pregnancy [9–11]. Between the ages of 18 to 24 years, a subset of ALSPAC offspring were invited to participate in three different neuroimaging studies, including the ALSPAC Psychotic Experiences (PE) study, where participants were assessed for PE using the psychotic-like symptoms semi-structured interview (PLIKS) [12, 13], administered by trained psychologists. Those who were found to have one or more PE were invited to undergo scanning. The presence of PE was confirmed using the clinical criteria of the Schedule for Clinical Assessment in Neuropsychiatry (SCAN) [14] and excluded experiences occurring due to waking, falling asleep, fever or drug consumption [15]. Participants who did not exhibit any symptoms of PE were randomly selected to form the control group and undergo scanning.

All data were acquired at Cardiff University Brain Research Imaging Centre (CUBRIC) on a 3 Tesla General Electric HDx (*GE Medical Systems*, Milwaukee, WI) using an eight-channel receive only head RF head coil T_1_-weighted structural images were acquired with a 3D fast spoiled gradient echo (FSPGR) sequence (TR = 7.8 ms, TE = 3.0 ms, TI = 450 ms, flip angle = 20°, voxel size = 1 mm^3^ isomorphic). T_2_-weighted whole brain scans were acquired using a coronal TSE sequence (for 3T: TR = 10,000 ms, TE = 14 ms, FA = 149, Bandwidth = 193 and NEX = 1; for 1.5T: TR = 9000 ms, TE = 64 ms, FA = 180, Bandwidth = 149 and NEX = 2; voxel size same as for T_1_ scans). sMRI images were processed using the automated FreeSurfer 6.0.0 software package for brain imaging [8]. Ethical approval for this study was given by both the Cardiff University School of Psychology Ethics Committee and the ALSPAC Ethics and Law Committee, and informed consent was obtained from all participants (ALSPAC project ID B709).

##### ASRB – Australian Schizophrenia Research Bank

The ASRB is a comprehensive biobank of clinical, neuroimaging and genetic data acquired in individuals with schizophrenia and healthy comparison individuals [16]. Assessments consist of the Diagnostic Interview for Psychosis (DIP), which is used to establish a lifetime diagnosis of a psychotic disorder, as well as present and lifetime substance use disorder diagnoses, according to DSM-IV and ICD-10 criteria [17]. Participants are English speaking (required for neuropsychological assessments) and aged 18-65 years. Exclusion criteria included any neurological disorder, history of brain trauma followed by a long period of amnesia (> 24 h), intellectual disability (full-scale IQ < 70), movement disorders, current substance dependence, as well as electroconvulsive therapy in the past 6 months. Controls were participants who had not personal or family history of psychosis or bipolar 1 disorder.

Participants were recruited from five sites in Australia, with all sites implementing the same recruitment procedures and MRI acquisition protocols. An individual travelled to all five sites and was scanned at each site to quantify gross inter-site differences. Structural and diffusion-weighted MRI scans of brain anatomy were acquired using *Siemens Avanto* 1.5T MRI scanners located in Melbourne, Sydney, Brisbane, Perth and Newcastle. A Siemens MRI phantom was scanned at each site to enable inter-site calibration and minimize potential inter-site variability. The same acquisition sequence was used at all sites. Structural T_1_-weighted images were acquired using an optimized MPRAGE sequence (TR = 1980 ms, TE = 4.3 ms, flip angle = 15, FOV = 256x256x176 mm, voxel resolution = 1 mm^3^ isotropic) [18]. Participants showing gross artefacts, cerebellar cropping and/or significant head motion were excluded, following protocols established as part of a prior study in this cohort. Calculation was undertaken using FreeSurfer 6.0.1. Approval to contribute ASRB data to this study was granted by the ASRB Access Committee on December 17, 2020.

##### B-SNIP – Bipolar & Schizophrenia Consortium for Parsing Intermediate Phenotypes

B-SNIP is a multicenter dataset available through NDAR (ID: 2274) that includes individuals diagnosed with psychosis, including schizophrenia (SCZ), schizoaffective disorder (SAD) and psychotic bipolar disorder (BDP); as well as their first-degree relatives and healthy volunteers without psychotic illness in their immediate family [19, 20]. Each proband was rated on a Schizo-Bipolar Scale, developed by the B-SNIP collaboration, which graded their component of the DSM’s criteria for each of the three psychosis disorders. The Schizo-Bipolar scale is graded from 1 (bipolar-like) to 10 (schizophrenia-like) with SAD in the middle. Volunteers participated in a SCID interview and relatives participated in additional interviews. Diagnosis was made in a consensus conference with multiple study-trained clinicians. Family history data were collected at a minimum from the most informed family member; more detailed information was gathered whenever possible.

Whole-brain structural MRI three-dimensional acquisitions were performed on 3T scanners (*GE Signa*, *Philips Achieva*, *Siemens Allegra*, and *Siemens Trio*) [21]. All participants at each site were scanned on the same magnet. High resolution isotropic T_1_-weighted MP-RAGE sequences were obtained following the Alzheimer’s Disease Neuroimaging Initiative (ADNI) protocol (<https://adni.loni.usc.edu/methods/mri-tool/mri-analysis/>). For the present study raw structural T_1_-weighted scans were processed with FreeSurfer 6.0.1.

##### LA5c – UCLA Consortium for Neuropsychiatric Phenomics LA5c Study

The LA5c dataset comprises data on OpenNeuro from the UCLA Consortium for Neuropsychiatric Phenomics LA5c Study (<https://openfmri.org/dataset/ds000030/>) which includes imaging of a large group of healthy individuals from the community as well as samples of individuals diagnosed with schizophrenia, bipolar disorder, and ADHD. To be included individuals had to be either “White, Not of Hispanic or Latino Origin” or “Hispanic or Latino, of Any Race” following NIH designations of racial and ethnic minority groups and have completed at least 8 years of education (other racial and ethnic minority groups were excluded because this was thought to increase risk of confounding planned genetic studies). The participants, ages 21-50, were recruited by community advertisements from the Los Angeles area and completed extensive neuropsychological testing, in addition to fMRI and T_1_-weighted Anatomical MPRAGE scanning.

All participants were scanned on a 3T *Siemens Trio* at a single-center [22]. T_1_-weighted MP-RAGE images were acquired with FOV = 250, 256x256 matrix, 176 1.0 mm sagittal partitions, TI = 1.1 s, TE = 3.5-3.3 ms, TR = 2.53 s and flip angle = 7. For the present study T_1_-weighted scans were processed with FreeSurfer 6.0.1.

##### MCIC – Mental Illness and Neuroscience Discovery (MIND) Institute Clinical Imaging Consortium

The Mental Illness and Neuroscience Discovery Institute (MIND) Institute, now the Mind Research Network (MRN; <https://www.mrn.org/>) formed the MIND Clinical Imaging Consortium (MCIC) in 2003 to conduct a multi-institutional, cross-sectional study of patients with schizophrenia and demographically matched, by sex and age, healthy controls to identify quantitative neuroimaging biomarkers for this devastating disease [23]. All subjects were between the ages of 18 and 60 and spoke English as their native language. To be included in the schizophrenia cohort, patients had to meet DSM-IV diagnostic criteria for schizophrenia, schizoaffective or schizophreniform disorder. Concerted effort was made to recruit patients early in the course of their illness and especially those who were antipsychotic drug-naïve. Patients were not excluded from the study because of substance abuse or dependence withing the past month, except for 6 patients who were found to meet criteria for current abuse after the study data was collected. The healthy control subjects without any current or previous psychiatric illness, including substance abuse or dependence, were matched within site to the patient cohort for age, sex, and parental education. Control subjects who had not been diagnosed with any psychiatric disorders, but had been medicated with antidepressants, anti-anxiety medication or medication for sleep disturbance were included in the study provided that the duration of their medication did not exceed 2 months of lifetime use and no medication was used within the 6 months preceding the baseline MRI scan. Both patients and controls were excluded if they had (1) an IQ < 70 based on a standardized IQ test, (2) history of a head injury resulting in prolonged loss of consciousness, neurosurgical procedure, neurological disease, history of skull fracture, severe or disabling medical conditions, or (3) a contraindication for MRI scanning such as pregnancy, metal in body or head including implanted pacemaker, medication pump, vagal stimulator, deep brain stimulator, implanted TENS unit, or ventriculoperitoneal shunt.

For each subject (patients and healthy controls), structural, DWI, and functional MR data were collected (*Siemens Sonata*, *Siemens Trio*, and *GE Signa*). Standardization of acquisition across sites was previously evaluated in a separate calibration and validation study [24]. All imaging data were collected using scanners with field strengths of 1.5T or 3.0T and using closely matched acquisition sequences [23]. Coronal T_1_ and T_2_ scans were collected during each structural imaging session. Imaging parameters for the T_1_ scans were, for 3T: TR = 2530 ms, TE = 3.79 ms, FA = 7, TI = 1100 and Bandwidth = 181; for 1.5T: TR = 12 ms, TE = 4.76 ms, FA = 20 and Bandwidth = 110; 0.625×0.625 mm voxel size; slice thickness 1.5 mm; FOV 256×256×128 cm matrix; FOV = 16 cm (could be increased to 18 cm when needed for full brain coverage). T_1_- weighted scans took approximately 13 minutes each and 2-3 volumes were acquired. T_2_-weighted whole brain scans were acquired using a coronal TSE sequence with the following parameters for 3T: TR = 10,000 ms, TE = 14 ms, FA = 149, Bandwidth = 193 and NEX = 1; for 1.5T: TR = 9000 ms, TE = 64 ms, FA = 180, Bandwidth = 149 and NEX = 2; voxel size same as for T_1_ scans. Acquisition of each volume took approximately 14.5 minutes, with 2 volumes acquired. Structural MRI data from the T_1_-weighted volumes were also co-registered, motion corrected, averaged, and analyzed in an automated manner with atlas-based FreeSurfer 6.0.1 software. The MCIC imaging and clinical data are available through the COINS (COllaborative Informatics Neuroimaging Suite) database [25] and may be freely used with no restrictions.

##### PAFIP – Programa de Atención a las Fases Iniciales de Psicosis

PAFIP subjects were screened for the following criteria [26, 27]: (1) age 15 to 60 years; (2) DSM-IV criteria for a principal diagnosis of schizophreniform disorder, schizophrenia, schizoaffective disorder, brief reactive psychosis, schizotypal personality disorder, or psychosis not otherwise specified; (3) habitually living in the catchment area; (4) no prior treatment with antipsychotic medication or, if previously treated, a total lifetime of adequate antipsychotic treatment of less than 6 weeks. Patients were excluded for any of the following reasons: 1) met the DSM-IV criteria for drug dependence (except nicotine dependence), 2) met the DSM-IV criteria for mental retardation, or 3) had a history of neurological disease or head injury. There were no additional exclusion criteria for MRI except those specific to scanning logistics (e.g., claustrophobia, braces). The healthy volunteer group was recruited from the community through advertisements. They were required to have no current or previous psychiatric, mental retardation, neurological or general medical illnesses, including substance dependence and significant loss of consciousness, as determined by using an abbreviated version of the Comprehensive Assessment of Symptoms and History (CASH) [28]. They were selected to have a similar distribution in age, gender, and drug use history to the patients. Clinical records and family interviews also confirmed the absence of psychosis in first-degree relatives. After a detailed description of the study, each subject gave written informed consent to participate.

Structural brain MRI scans were obtained using a 1.5 T General Electric SIGNA System (GE Medical Systems, Milwaukee, WI) and a 3 T Philips Medical Systems MRI scanner (Achieva, Best, The Netherlands) at the *Hospital Universitario Marqués de Valdecilla* (HUMV). The parameters for 1.5T were: TE = 5 ms, TR =24 ms, NEX = 2, rotation angle = 45°, FOV = 26×19.5 cm, slice thickness = 1.5 mm, and a matrix of 256×192, whereas for 3T were: TE = 3.7 ms, TR = 8.2 ms, flip angle = 8°, acquisition matrix = 256×256, voxel size = 0.94x0.94x1 mm and 160 contiguous slices [29]. T_1_-weighted images were first visually inspected for artifacts and gross anatomical abnormalities. Scan images were processed using the FreeSurfer 6.0.0 longitudinal stream.

##### UKB - UK Biobank

The UK Biobank (<https://www.ukbiobank.ac.uk/>) is a prospective cohort of 500,000 individuals from across the UK, aged between 40 and 69 at recruitment [30–32]. Of these individuals, 100,000 will undergo brain scanning [33]. Participants were excluded from the MRI study on the basis of standard MRI safety criteria such as metal implants, recent surgery, or conditions problematic for scanning such as hearing problems, breathing problems, or claustrophobia [34].

Brain imaging is being conducted on the 3T *Siemens Skyra* (software platform VD13) system using a 32-channel receive head coil. The T_1_ structural protocol is acquired at 1mm isotropic resolution (208x256x256) using a three-dimensional (3D) MPRAGE acquisition (R = 2, TI/TR = 800/2000 ms). Images were segmented into tissue types (gray matter, white matter, and cerebrospinal fluid) [35]. Cortical gray matter volume was estimated, comparing the segmented gray matter to an atlas reference (where the external skull surface is used to normalize for head size) [36]. The T_2_ protocol used a fluid-attenuated inversion recovery (FLAIR) contrast with the 3D SPACE optimized readout (R =2, PF 7/8, fat sat, TI/TR = 1800/5000 ms, elliptical). All T_1_-weighted images were processed using FreeSurfer 6.0.1. Further protocol details are available at <https://biobank.ctsu.ox.ac.uk/crystal/refer.cgi?id=2367>.

Ethical procedures for the UK Biobank are controlled by the Ethics and Guidance council (<http://www.ukbiobank.ac.uk/ethics>), and the study was conducted in accordance with the UK Biobank an Ethics and Governance Framework document (<https://www.ukbiobank.ac.uk/media/0xsbmfmw/egf.pdf>), with institutional review board approval by the North West Multi-center Research Ethics Committee.

#### **Centile-based normative model**

The out-of-sample centile method used here enabled us to calculate centiles without the need to re-train the model on the ~120,000 subjects used for the original normative curves [37]. Briefly, our out-of-sample data is aligned to the corresponding epoch of the normative trajectory, using maximum likelihood to estimate the study specific offsets (random effects) for three moments of the underlying statistical distributions: mean (μ), variance (σ), and skewness (ν) in an age- and sex-specific manner. This allows to compute centiles for each scan in a new study, on the same scale as the reference population curve, while accounting for study-specific ‘batch effects’.

#### **Spin test**

The spin test projects brain regions to a sphere using spherical coordinates generated during cortical-surface extraction. These spherical projections of brain annotation maps are rotated to randomize the relationship between cortical attributes and annotations [38, 39]. Each coordinate is assigned to its nearest rotated counterpart, resulting in a map where spatial autocorrelation is preserved, while the correspondence between parcels and annotations is randomized. Parcels that were closest to the medial wall were assigned the value of the nearest neighboring parcel instead. This procedure was performed at parcel resolution, rather than the vertex resolution, to prevent data upsampling, and it was repeated 10,000 times to generate a parcellation-specific rotation matrix (available at <https://github.com/frantisekvasa/rotate_parcellation>).

### Supplemental Figures

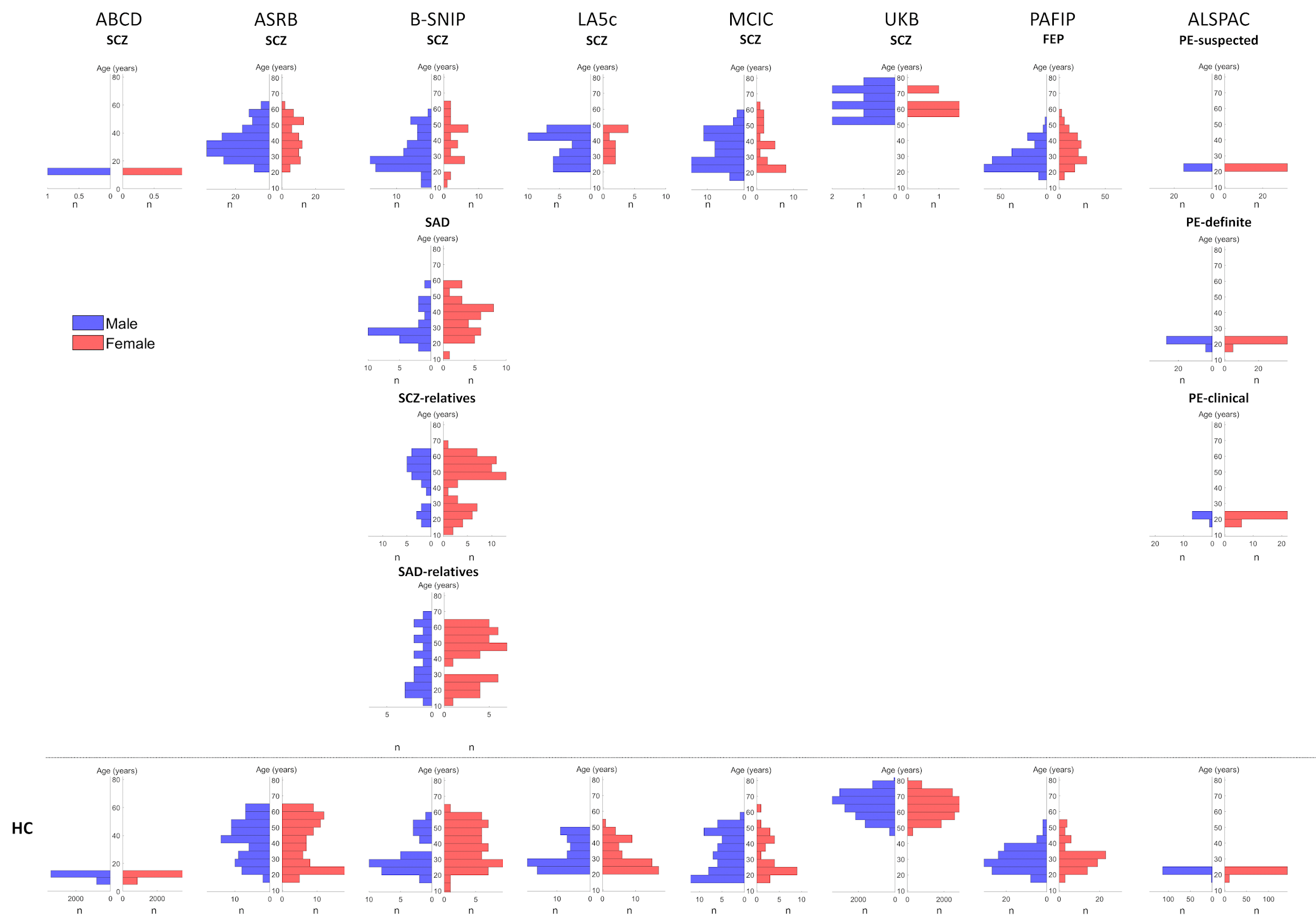

**Supplementary Fig. 1 Age distributions stratified by sex, diagnosis, and dataset.** The sample size (n) represents the number of individuals with the same sex, diagnosis, and dataset within the same 10-year age range.

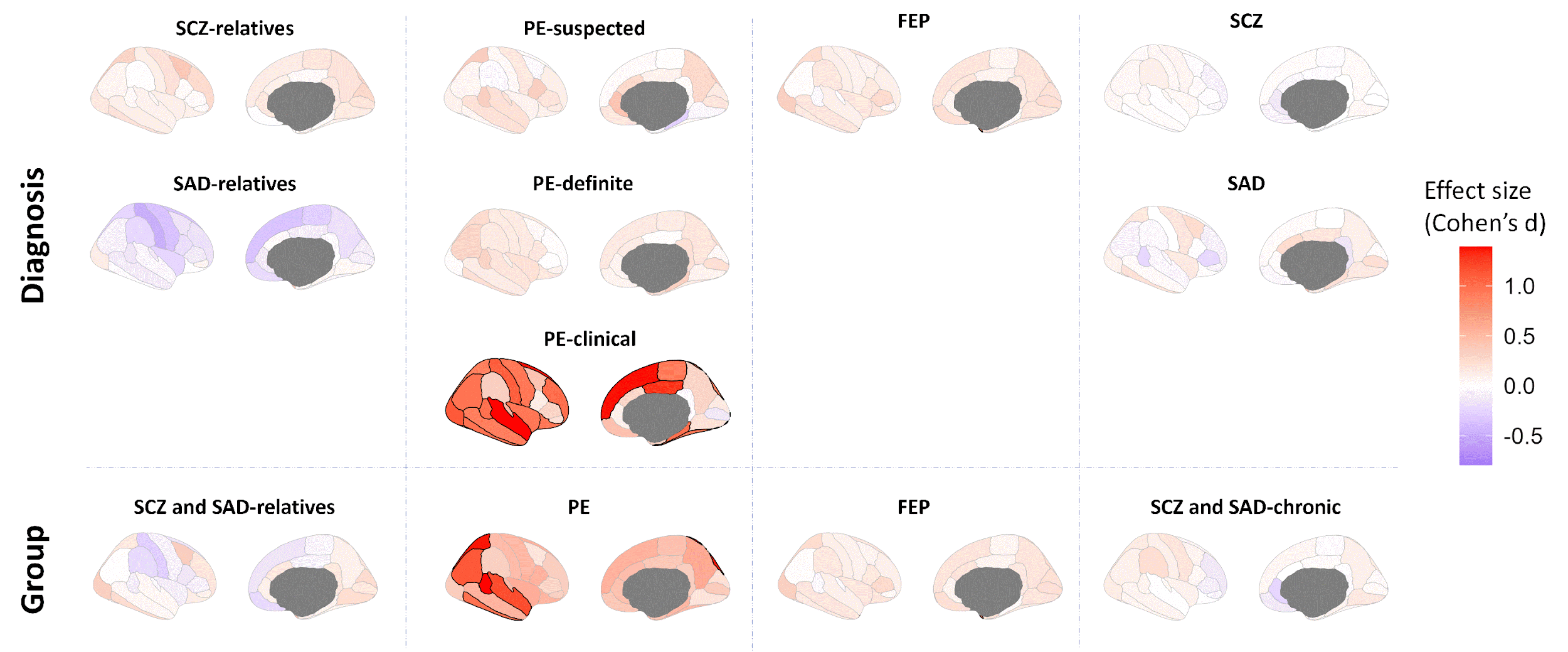

**Supplementary Fig. 2 Regional effect sizes of centiles between males and females.** Cohen’s d computed between regional centiles of males and females for each diagnosis and group to map the sex-effect on centiles. The highlighted regions show those regional effect sizes that exhibit significant differences between sexes after FDR correction (*P*_perm_ < 0.05). Positive values in red indicate a centile increase in males compared to females.

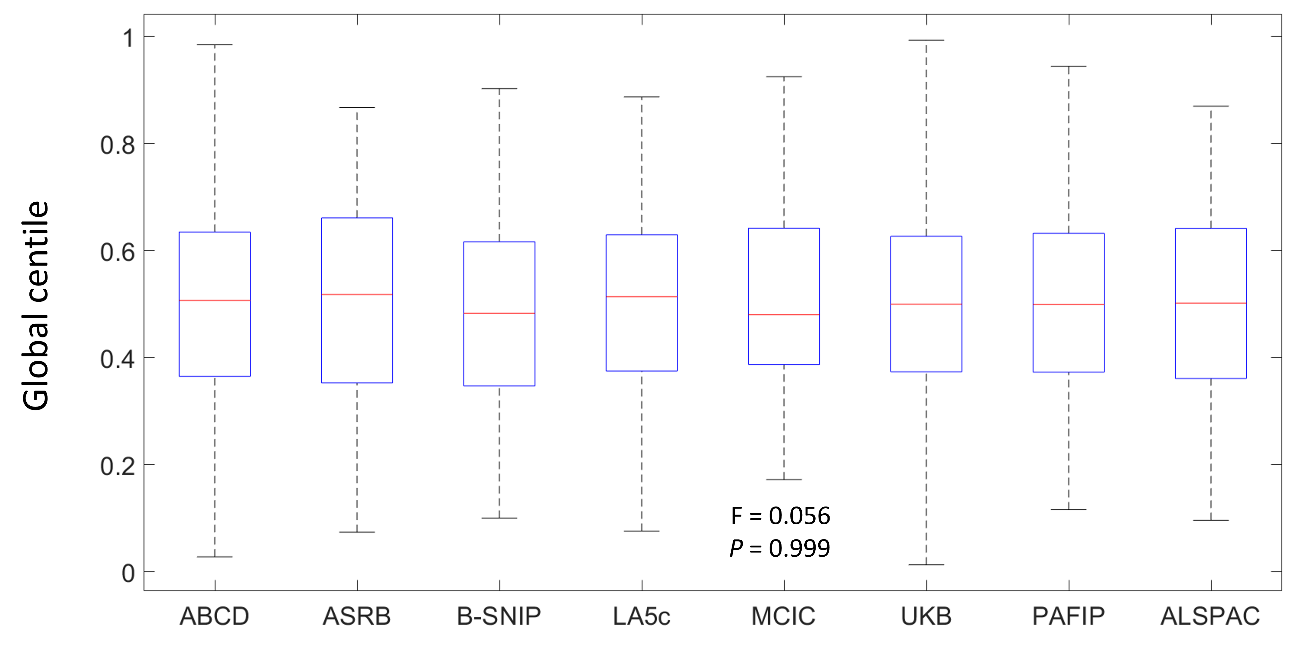

**Supplementary Fig. 3 Global HC centile distributions across studies.** Global HC centile distributions were computed for each dataset after averaging centiles across regions to represent the site-effect on centiles.

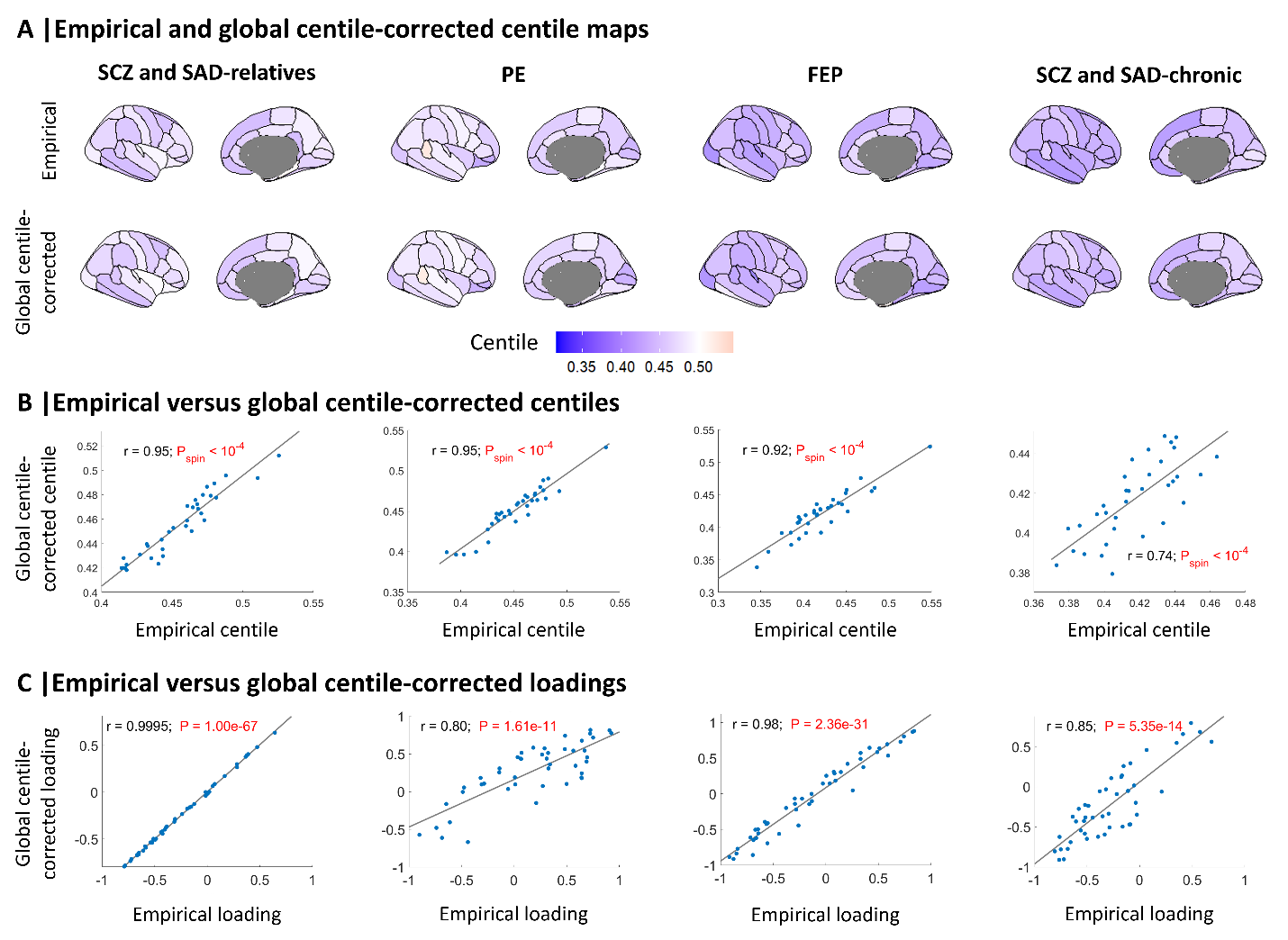

**Supplementary Fig. 4 Association between empirical regional centiles and global centile-corrected centiles.** (**A**) Maps of empirical MRI-derived centiles (top) and centiles after global-centile correction (bottom). (**B**) Correlation between empirical centiles and global centile-corrected centiles (*P*_spin_ < 0.05). (**C**) Correlation between empirical centiles and global centile-corrected neurobiological PCA-CCA loadings (*P*_spin_ < 0.05).

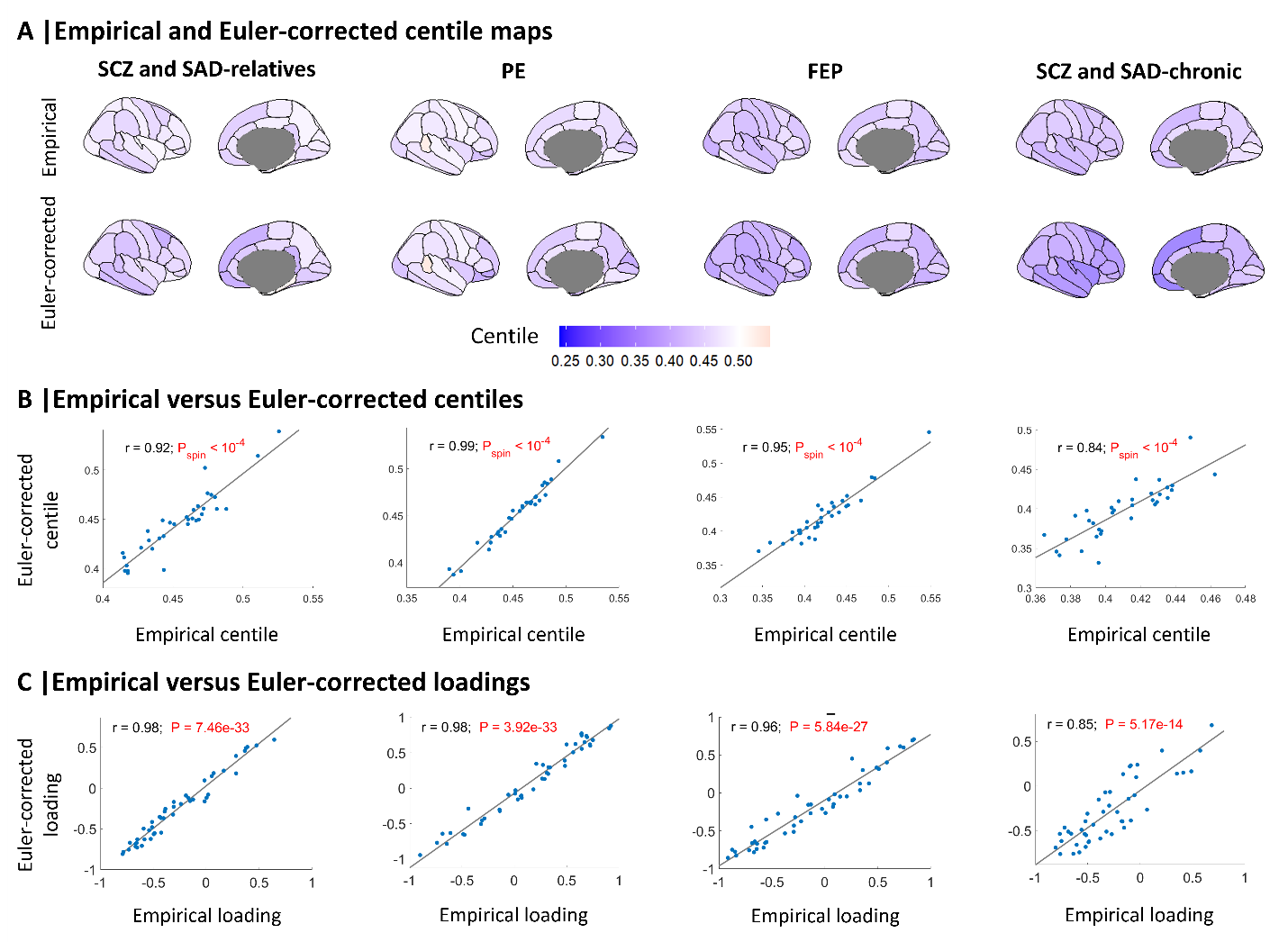

**Supplementary Fig. 5 Association between empirical regional centiles and Euler-corrected centiles.** (**A**) Maps of empirical MRI-derived centiles (top) and Euler-corrected centiles (bottom). (**B**) Correlation between empirical and Euler-corrected centiles (*P*_spin_ < 0.05). (**C**) Correlation between empirical and Euler-corrected neurobiological PCA-CCA loadings (*P*_spin_ < 0.05).

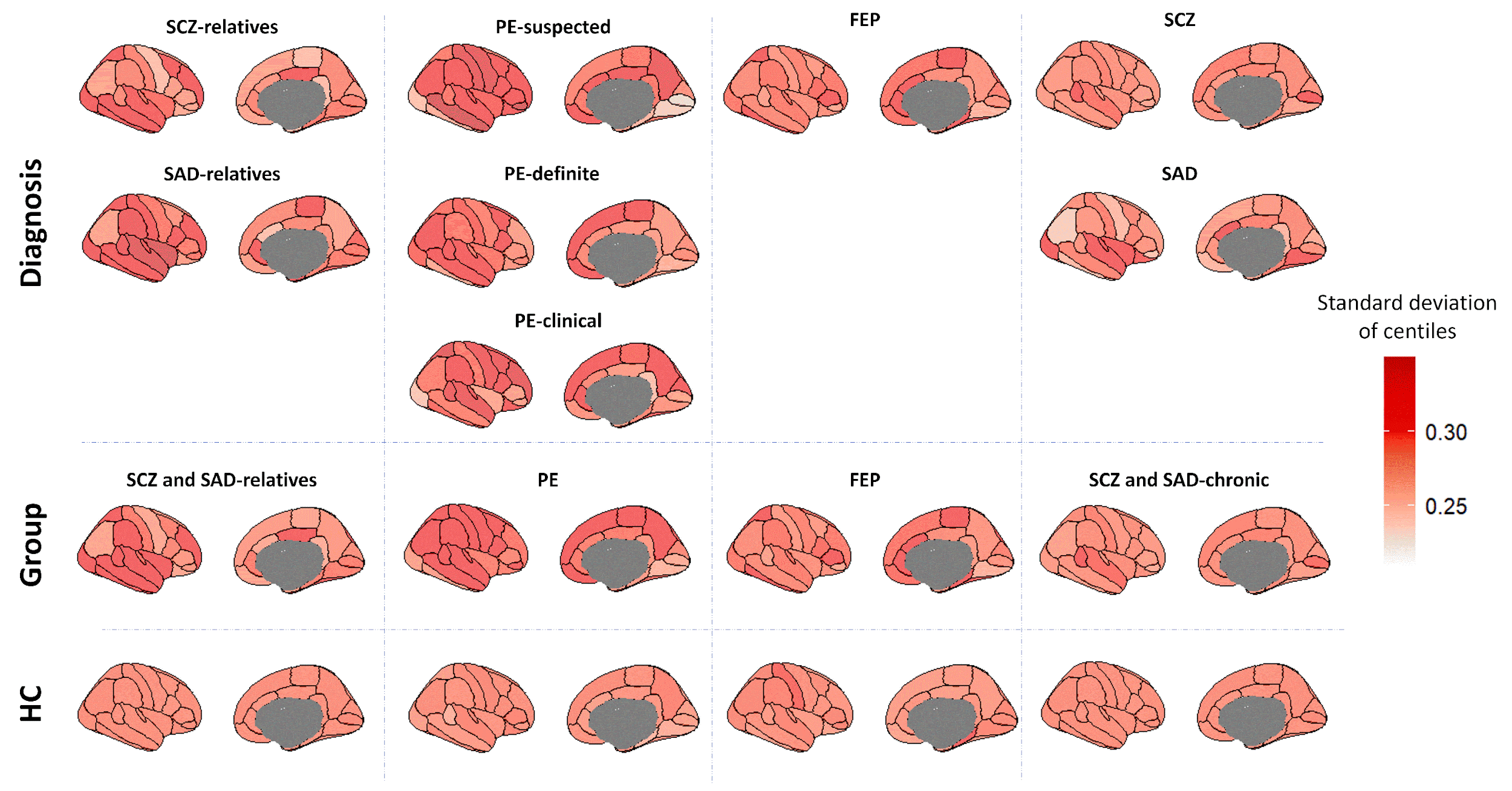

**Supplementary Fig. 6 Regional standard deviations of centiles.** Regional standard deviations of centiles were computed within each condition to generate a centile variability map for each diagnosis and group.

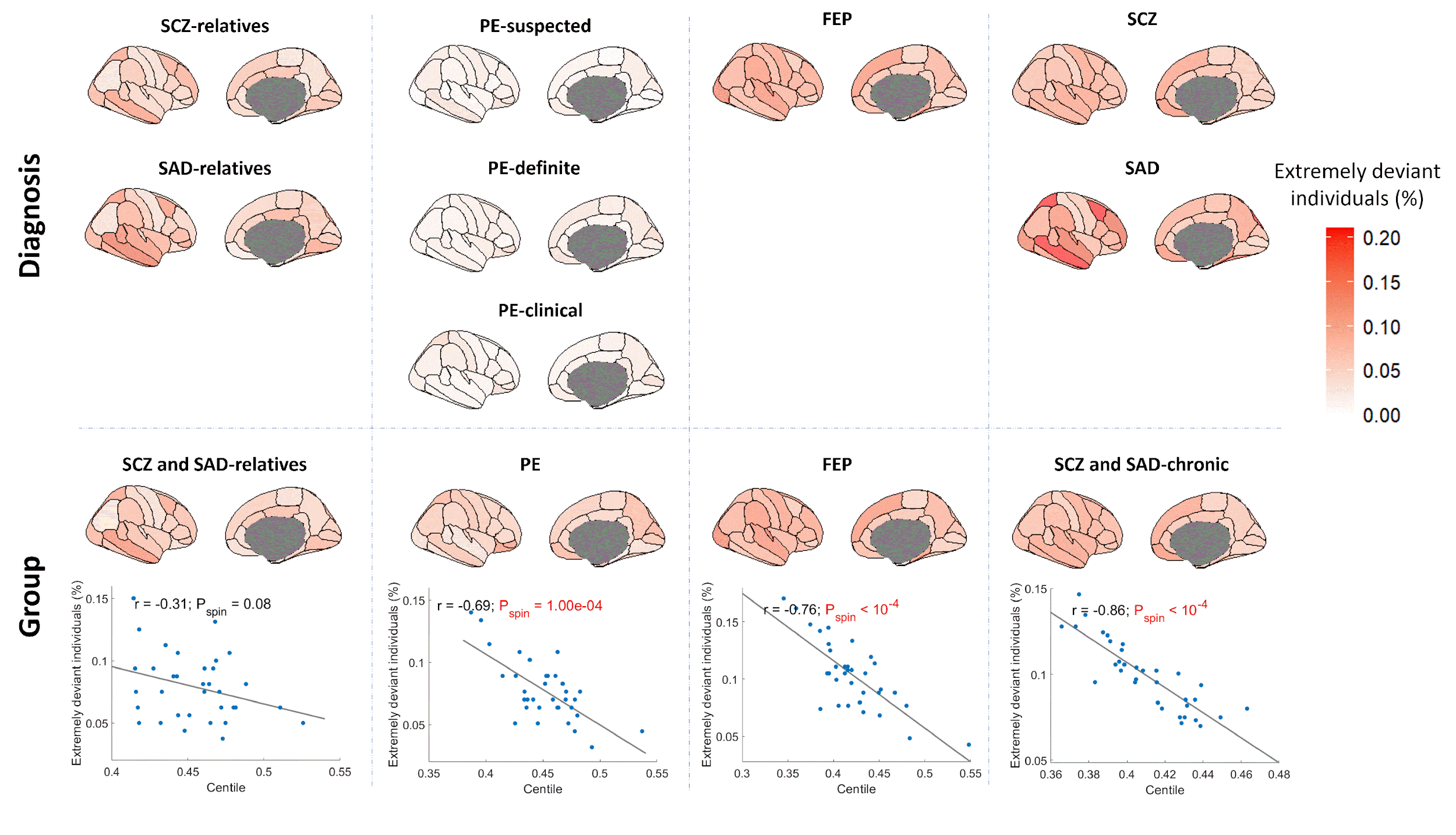

**Supplementary Fig. 7 Extremely deviant regions.** Extremely deviant regions were computed as the relative frequency of individuals (%) with extreme reductions in centile values (centile < 0.05). The association between extreme deviant regions and the centile maps represented in Figure 2 was computed for the groups to assess the consistency between approaches (*P*_spin_ < 0.05).

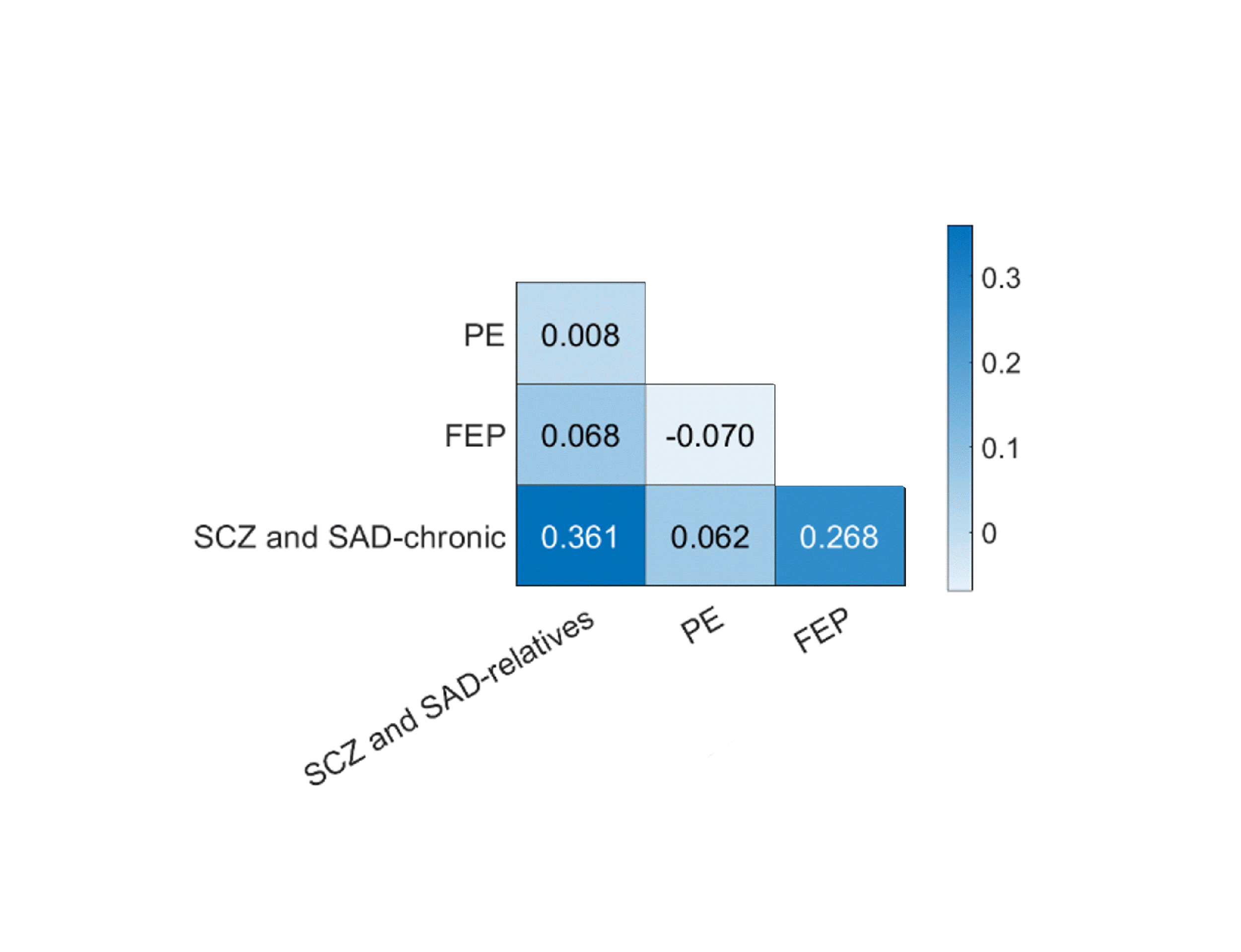

**Supplementary Fig. 8 Pearson correlation of centiles across regions between each pair of groups.** Significance was tested by using a permutation test in which group membership was randomly reassigned across the compared groups. No significant differences were observed in correlation among any pair of groups (all *P*_perm_ > 0.05).

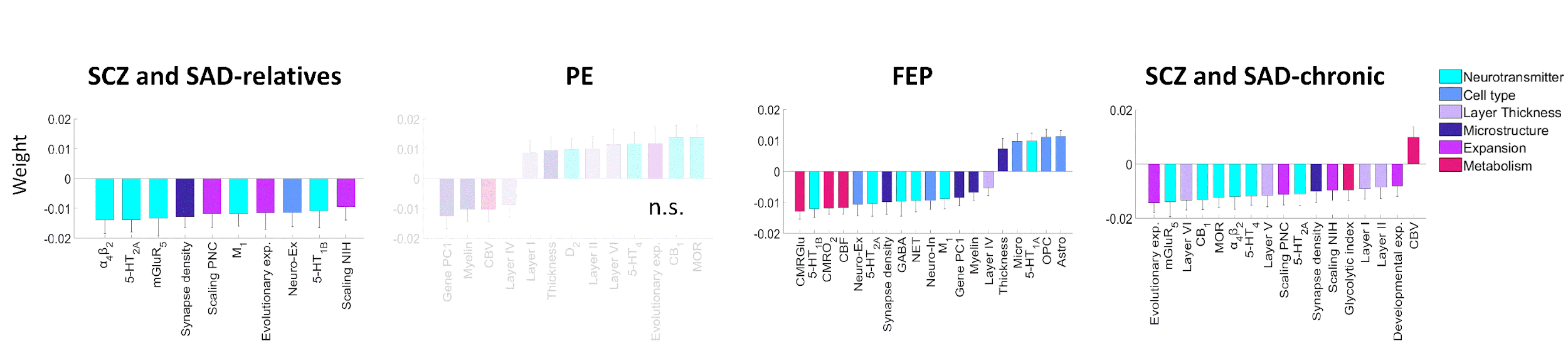

**Supplementary Fig. 9 PCA-CCA significant weights representing the contribution of each neurobiological feature to predicted centiles for each group.** Significant weights (*P*_spin_ < 0.05) were derived from neurobiological features using a PCA-CCA model for each group. Non-significant models are denoted as n.s (FDR-corrected *P*_spin_ > 0.05). Error bars represent the standard deviation.

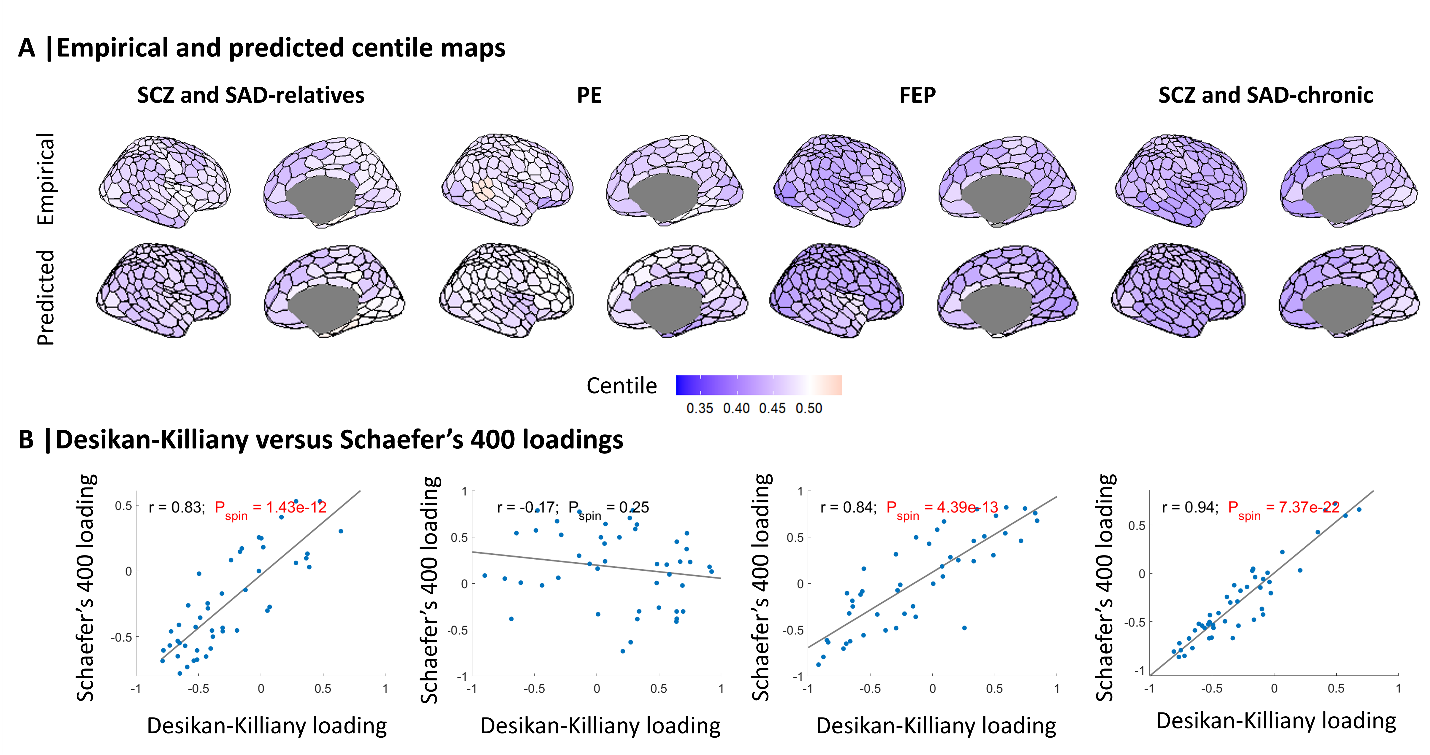

**Supplementary Fig. 10 Empirical and predicted centiles Schaefer’s 400-parcellated, and associated loadings from PCA-CCA models**. (**A**) Maps of empirical DK-parcellated centiles were matched to Schaefer’s 400 atlas (top) and their predicted PCA-CCA-derived centiles were obtained from neurobiological features (bottom). (**B**) Correlation between Schaefer’s 400 and DK loadings (*P*_spin_ < 0.05).

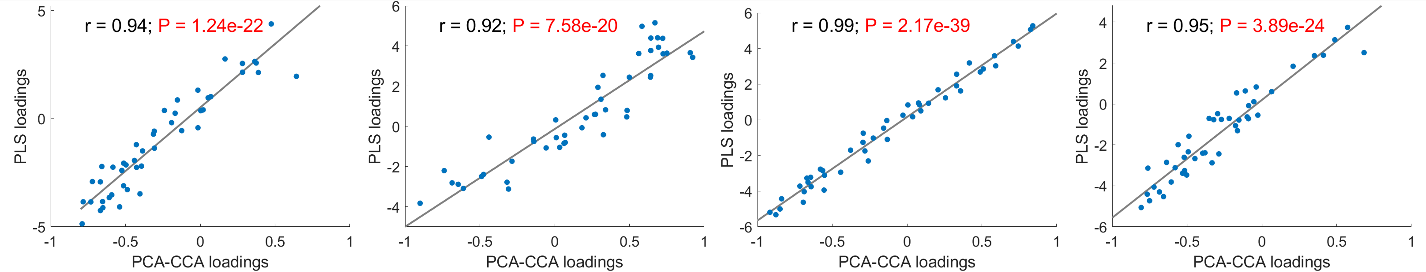

**Supplementary Fig. 11 Correlation between PLS- and PCA-CCA-derived loadings (*P*_spin_ < 0.05).**

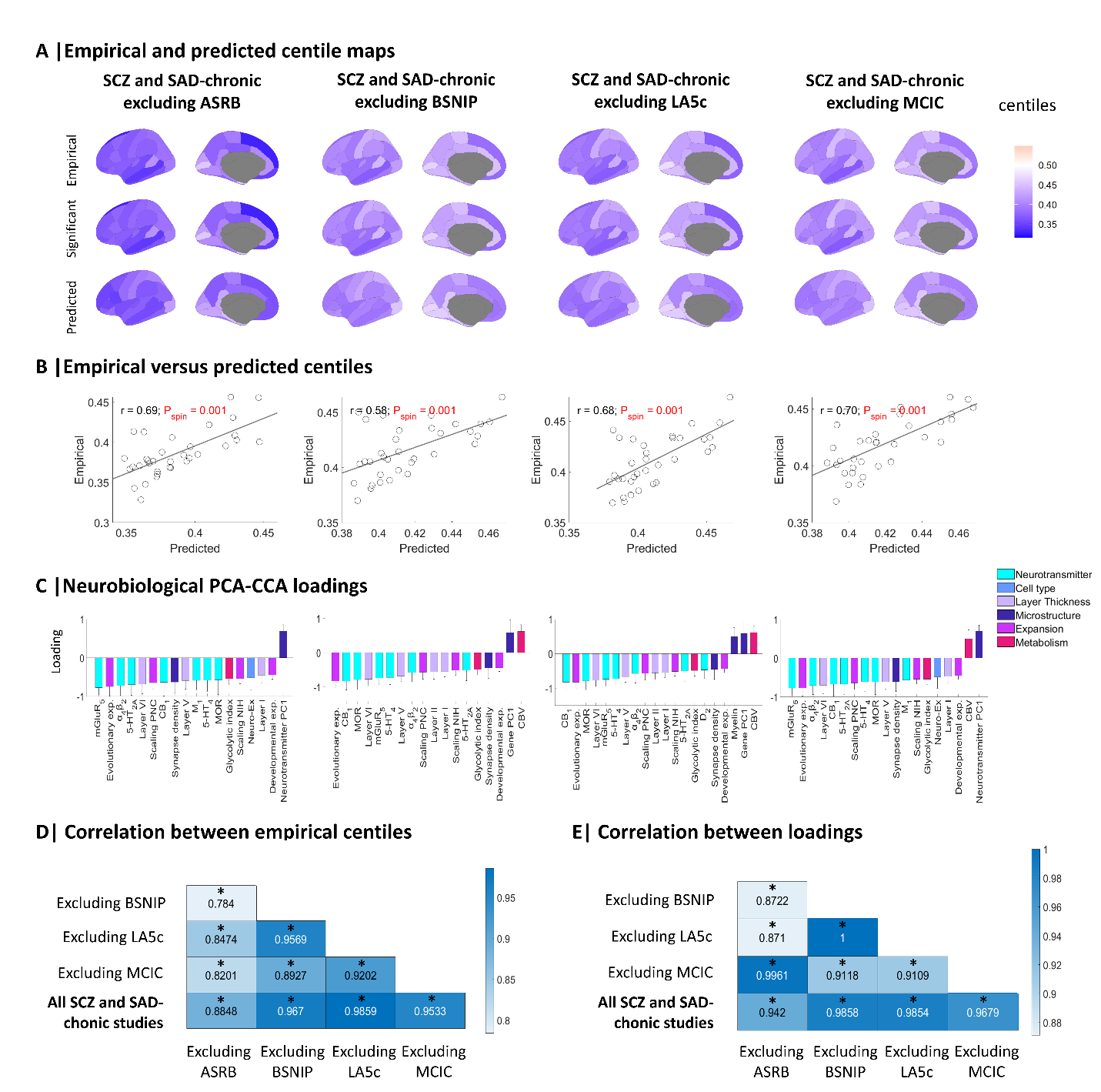

**Supplementary Fig. 12** **Empirical and predicted centiles, and associated loadings from PCA-CCA models of SCZ and SAD-chronic after leave-one-study-out cross-validation**. (**A**) Maps of empirical MRI-derived centiles (top), significant MRI-derived centiles (middle; Wilcoxon rank sum test FDR-corrected, *P* < 0.05), and predicted PCA-CCA-derived centiles from neurobiological features (bottom; FDR-corrected *P*_spin_ < 0.05). (**B**) Correlation between empirical and predicted regional centiles. (**C**) PCA-CCA significant loadings associated to each neurobiological map (*P*_spin_ < 0.05). Error bars represent the standard deviation. (**D**) Correlation between empirical centiles. Asterisks (*) indicate significant correlation of centiles between SCZ and SAD-chronic studies (FDR-corrected all *P*_spin_ < 10^-3^). (**E**) Correlation between loadings. Asterisks (*) indicate significant differences in correlation of loadings between SCZ and SAD-chronic studies (FDR-corrected *P*_Pearson_ < 10^-14^).

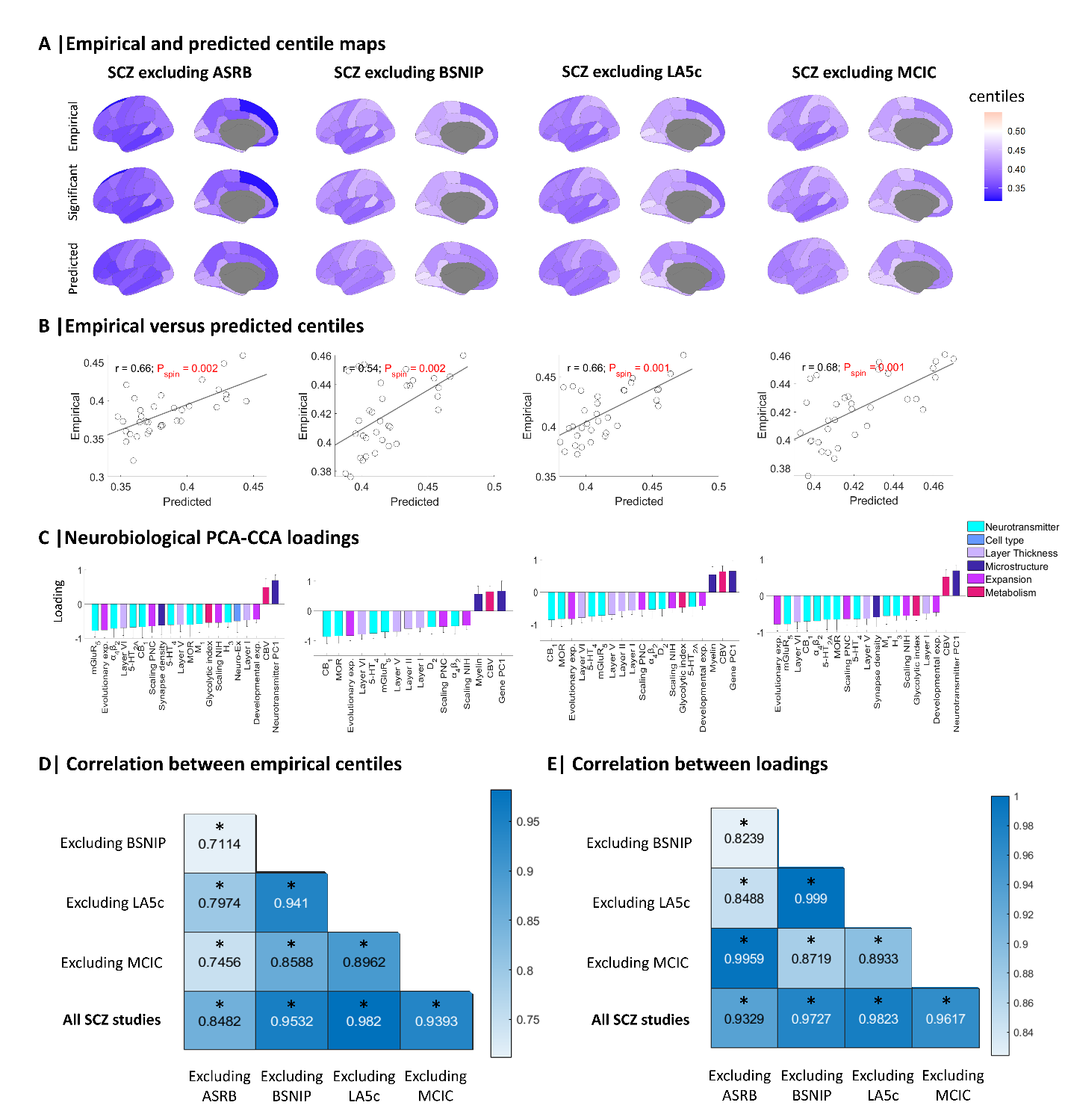

**Supplementary Fig. 13** **Empirical and predicted centiles, and associated loadings from PCA-CCA models of SCZ after leave-one-study-out cross-validation**. (**A**) Maps of empirical MRI-derived centiles (top), significant MRI-derived centiles (middle; Wilcoxon rank sum test FDR-corrected, *P* < 0.05), and predicted PCA-CCA-derived centiles from neurobiological features (bottom; FDR-corrected *P*_spin_ < 0.05). (**B**) Correlation between empirical and predicted regional centiles. (**C**) PCA-CCA significant loadings associated to each neurobiological map (*P*_spin_ < 0.05). Error bars represent the standard deviation. (**D**) Correlation between empirical centiles. Asterisks (*) indicate significant correlation of centiles between SCZ studies (FDR-corrected *P*_spin_ < 10^-3^). (**E**) Correlation between loadings. Asterisks (*) indicate significant differences in correlation of loadings between SCZ studies (FDR-corrected *P*_Pearson_ < 10^-11^).

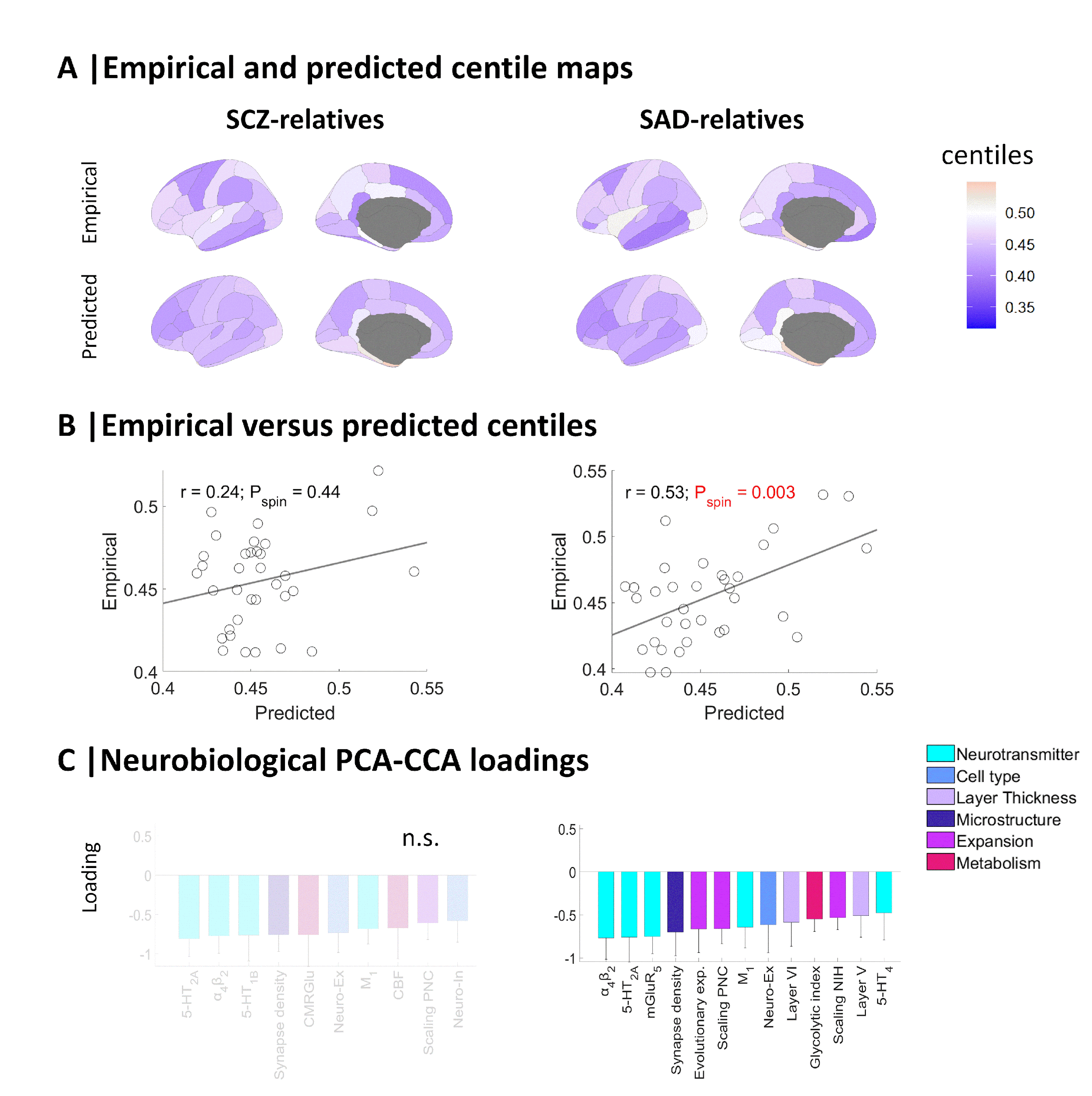

**Supplementary Fig. 14 Empirical and predicted centiles, and associated loadings from PCA-CCA models of SCZ and SAD relatives**. (**A**) Maps of empirical MRI-derived centiles (top) and predicted PCA-CCA-derived centiles from neurobiological features (bottom). (**B**) Correlation between empirical and predicted regional centiles. (**C**) PCA-CCA significant loadings associated to each neurobiological map (*P*_spin_ < 0.05). Non-significant models are denoted as n.s (FDR-corrected *P*_spin_ > 0.05). Error bars represent the standard deviation.

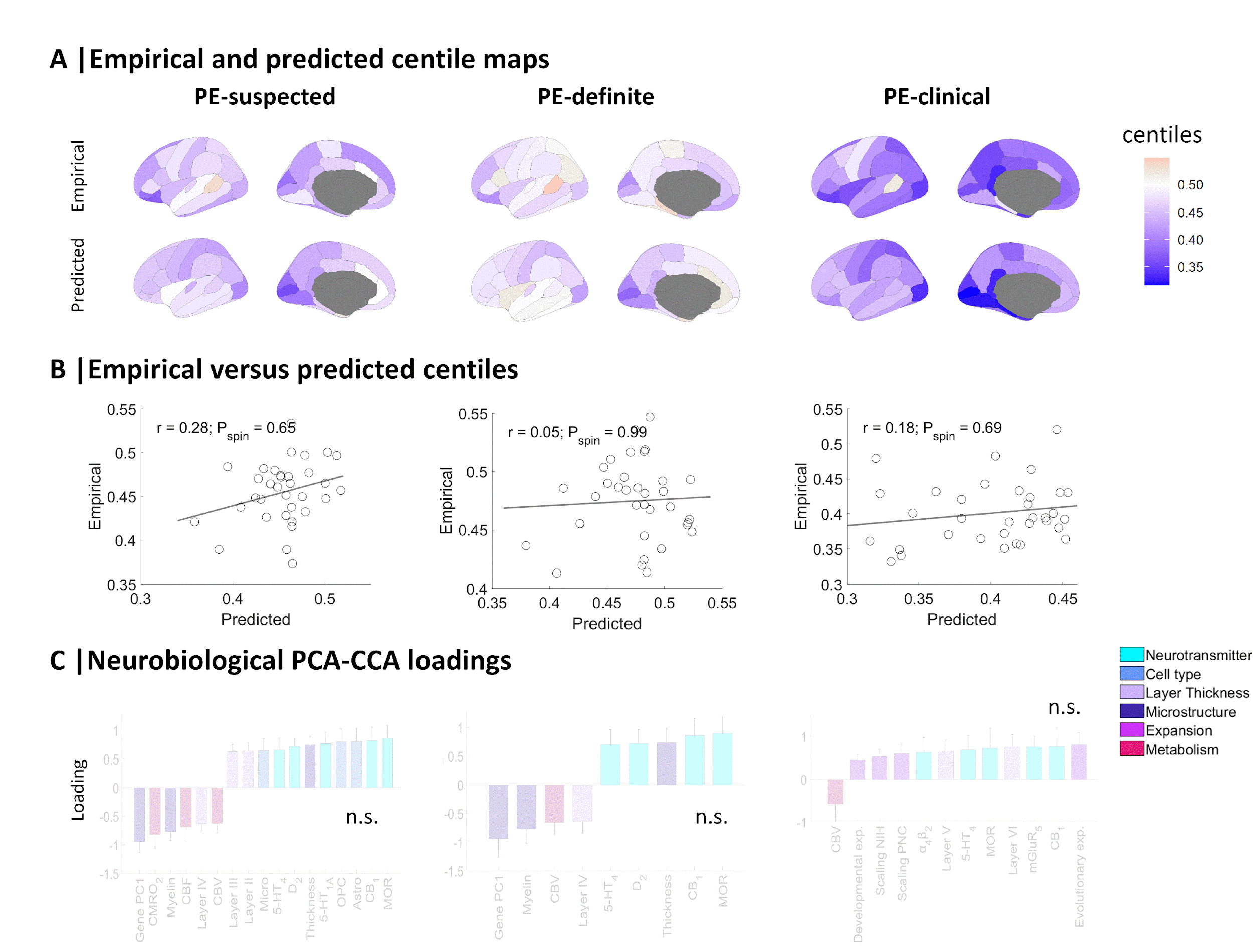

**Supplementary Fig. 15** **Empirical and predicted centiles, and associated loadings from PCA-CCA models of individuals who reported operationally-defined psychotic experiences, rated as ‘suspected’, ‘definite’, or ‘clinical’**. (**A**) Maps of empirical MRI-derived centiles (top) and predicted PCA-CCA-derived centiles from neurobiological features (bottom). (**B**) Correlation between empirical and predicted regional centiles. (**C**) PCA-CCA significant loadings associated to each neurobiological map (*P*_spin_ < 0.05). Non-significant models are denoted as n.s (FDR-corrected *P*_spin_ > 0.05). Error bars represent the standard deviation.

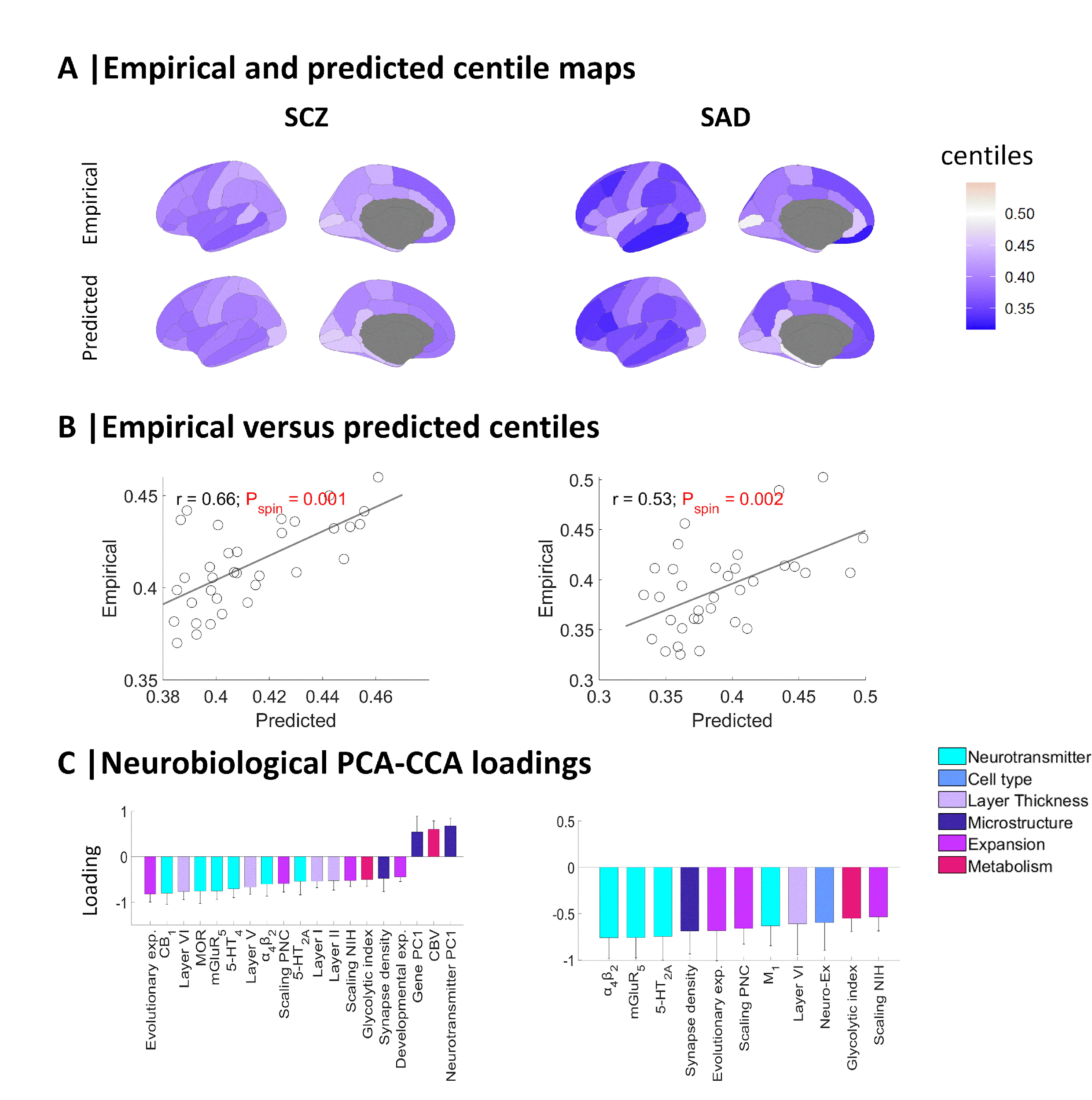

**Supplementary Fig. 16** **Empirical and predicted centiles, and associated loadings from PCA-CCA models of SCZ and SAD**. (**A**) Maps of empirical MRI-derived centiles (top) and predicted PCA-CCA-derived centiles from neurobiological features (bottom; FDR-corrected *P*_spin_ < 0.05). (**B**) Correlation between empirical and predicted regional centiles. (**C**) PCA-CCA significant loadings associated to each neurobiological map (*P*_spin_ < 0.05). Error bars represent the standard deviation.

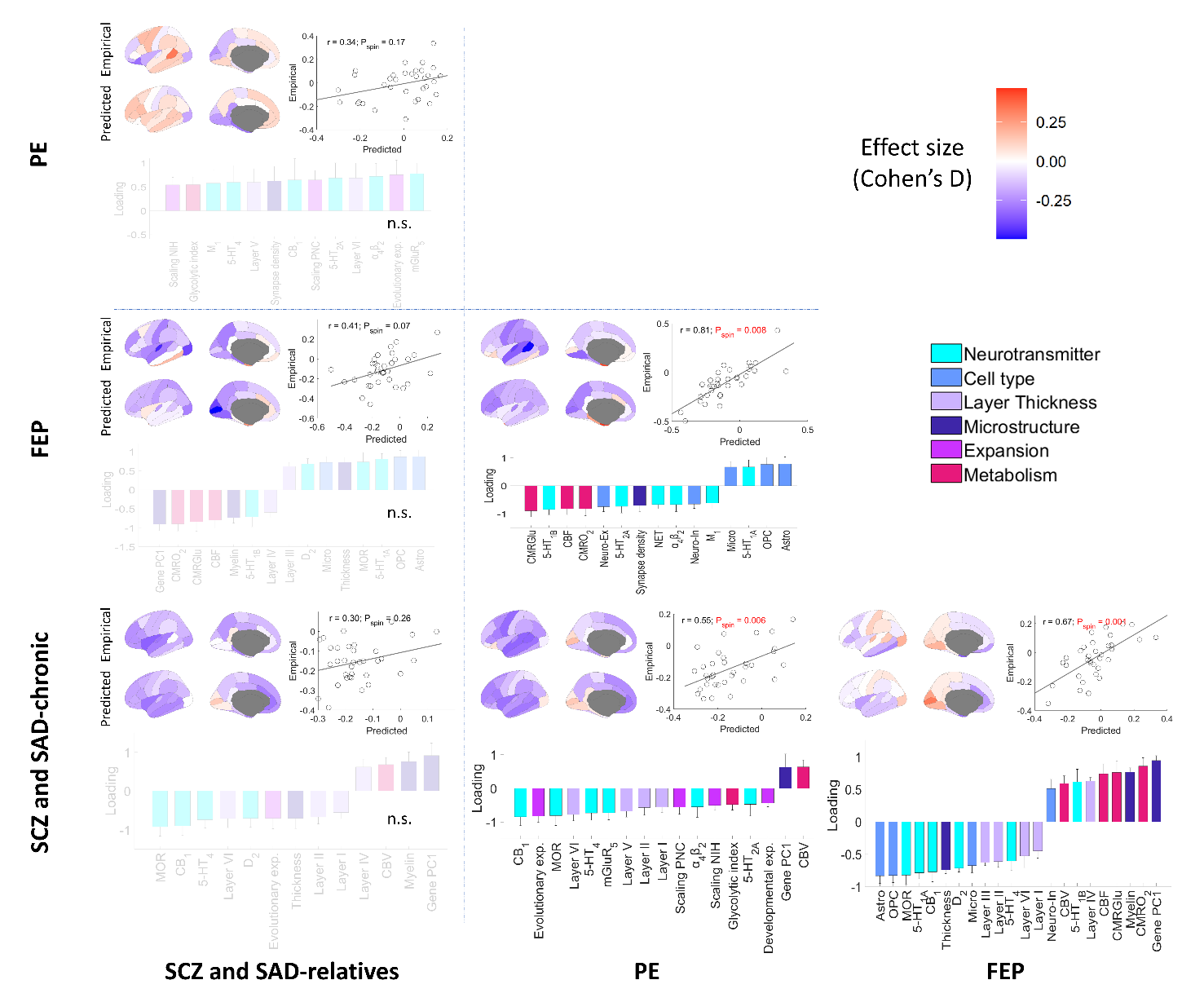

**Supplementary Fig. 17 Empirical and predicted effect sizes between pairs of psychosis-related groups, and associated loadings from PCA-CCA models.** Each panel illustrates the empirical and the PCA-CCA predicted effect sizes of centile maps (top-left); the correlation between empirical and predicted effect sizes (top-right); and the neurobiological PCA-CCA significant loadings (bottom; *P*_spin_ < 0.05). Non-significant models are denoted as n.s (FDR-corrected *P*_spin_ > 0.05). Error bars represent the standard deviation.

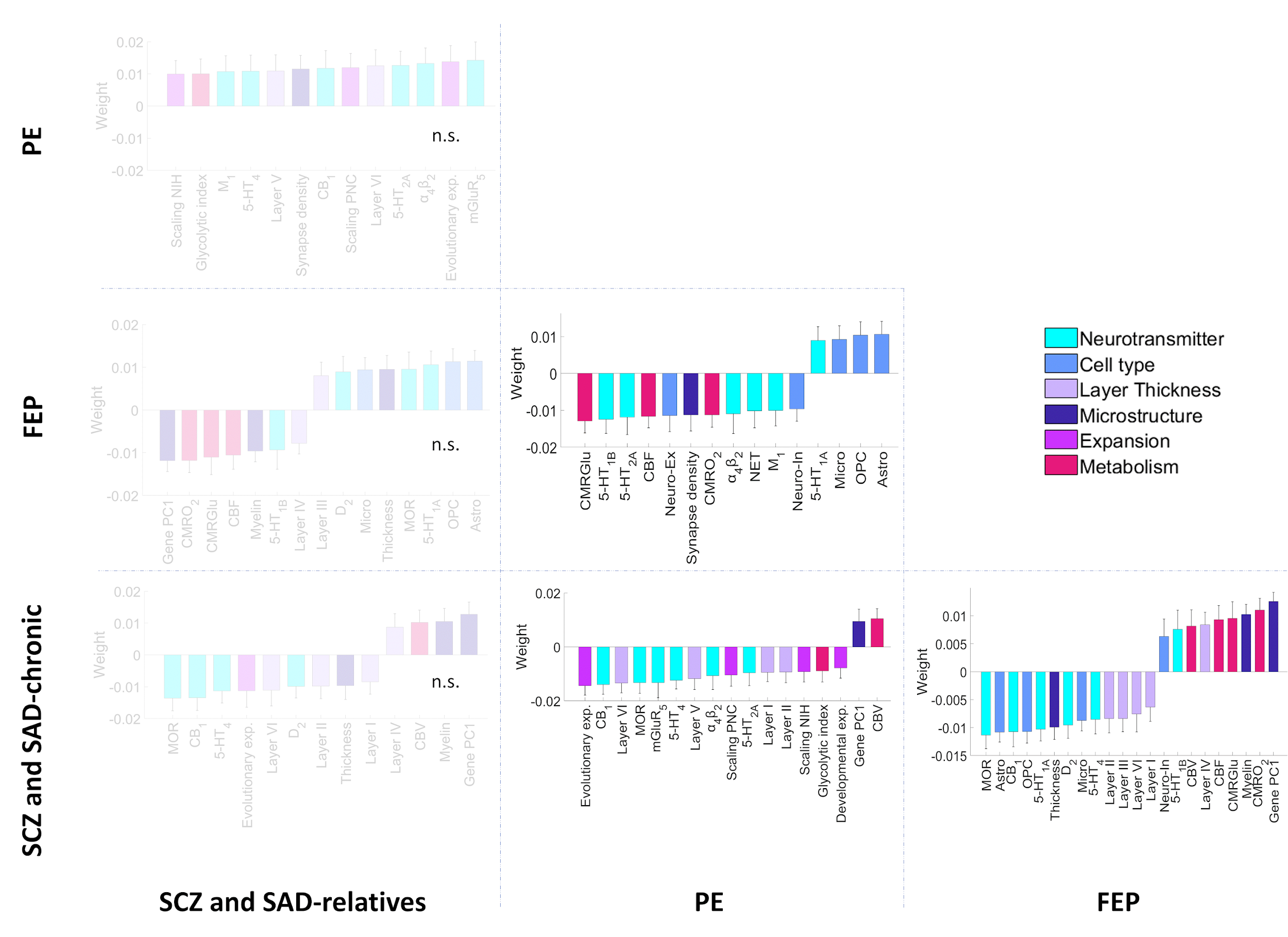

**Supplementary Fig. 18 PCA-CCA weights of each neurobiological feature contribution to the predicted effect sizes of centiles between groups.** Significant weights (*P*_spin_ < 0.05) were derived from neurobiological features using a PCA-CCA model for each pair of groups. Non-significant models are denoted as n.s (FDR-corrected *P*_spin_ > 0.05). Error bars represent the standard deviation.

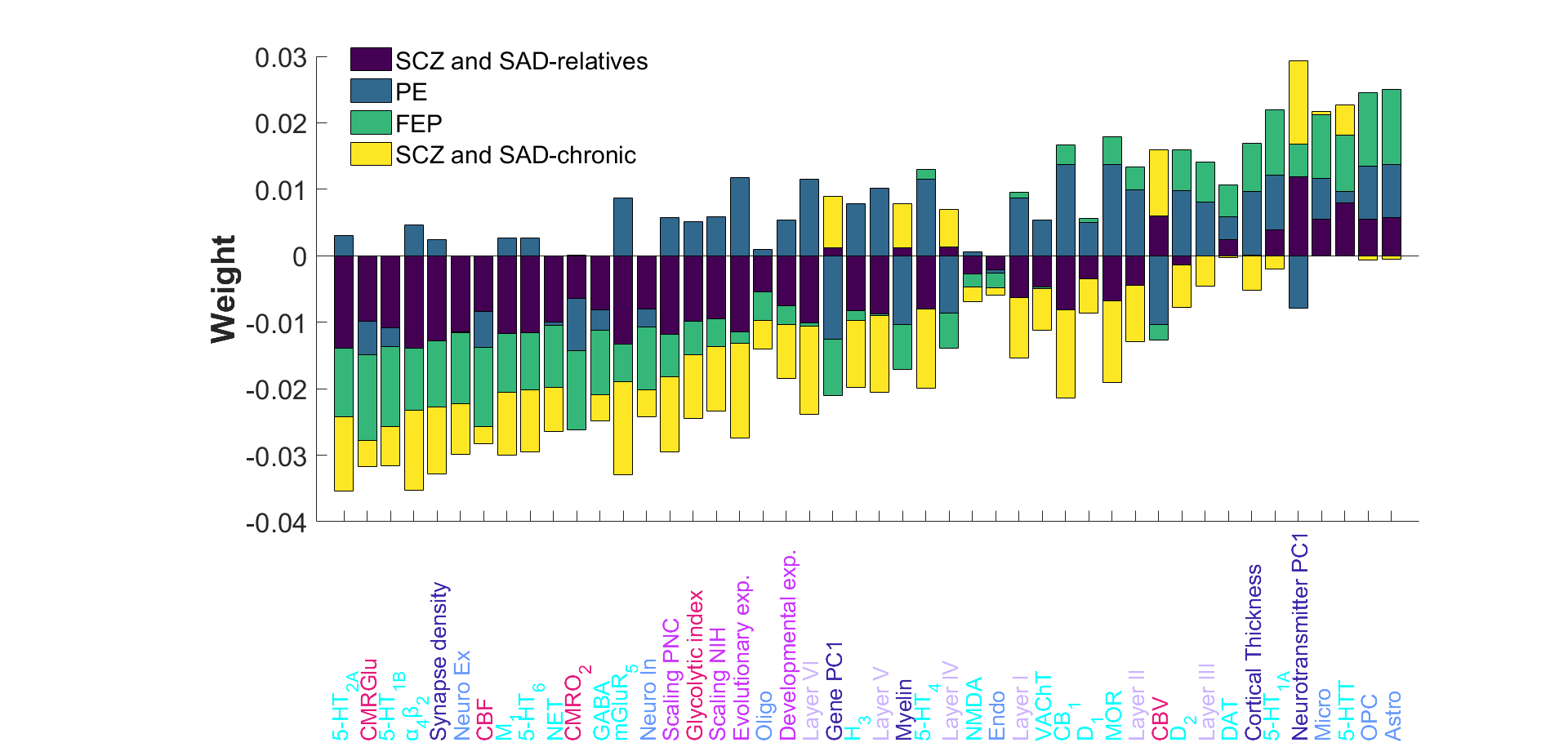

**Supplementary Fig. 19 Neurobiological PCA-CCA weights compared across psychosis-related groups.** Stacked neurobiological weights of each group, regardless of their significance, were ranked from the most negative to the most positive average contribution.

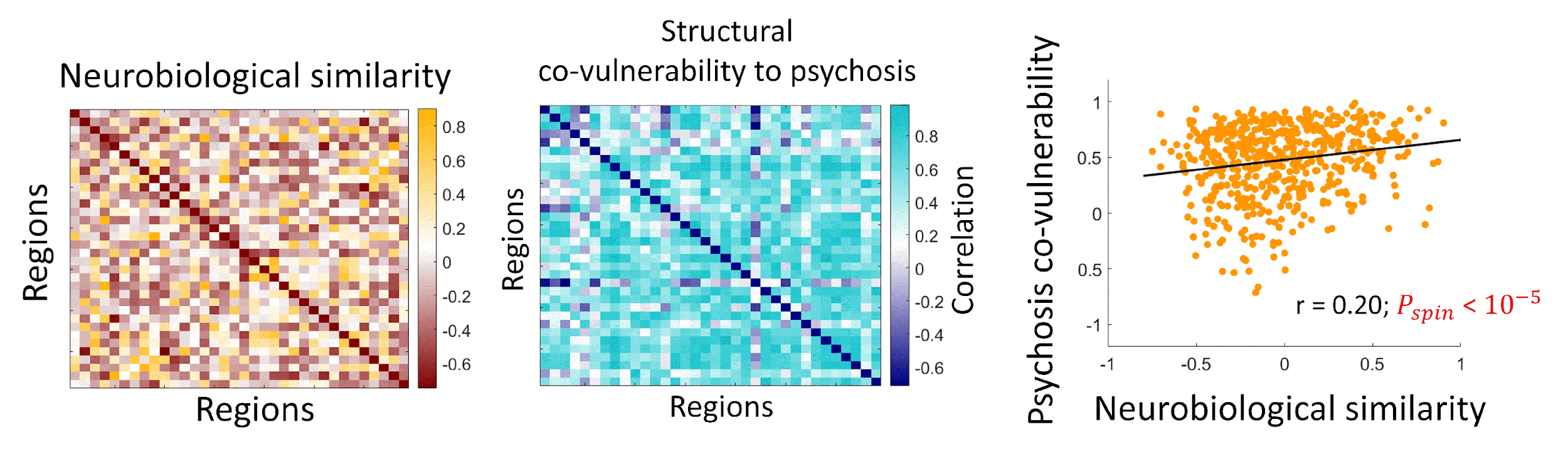

**Supplementary Fig. 20 Association between neurobiological similarity and structural co-vulnerability to diagnosis of psychosis.** Neurobiological similarity matrix obtained by correlating the regional patterns of neurobiological features in HC (left). Structural co-vulnerability to psychosis matrix constructed by correlating the regional patterns of the effect sizes of centiles across psychosis-related diagnoses (middle). Correlation between both matrices (right; Pearson’s correlation r = 0.20, *P*_spin_ < 10^-5^).

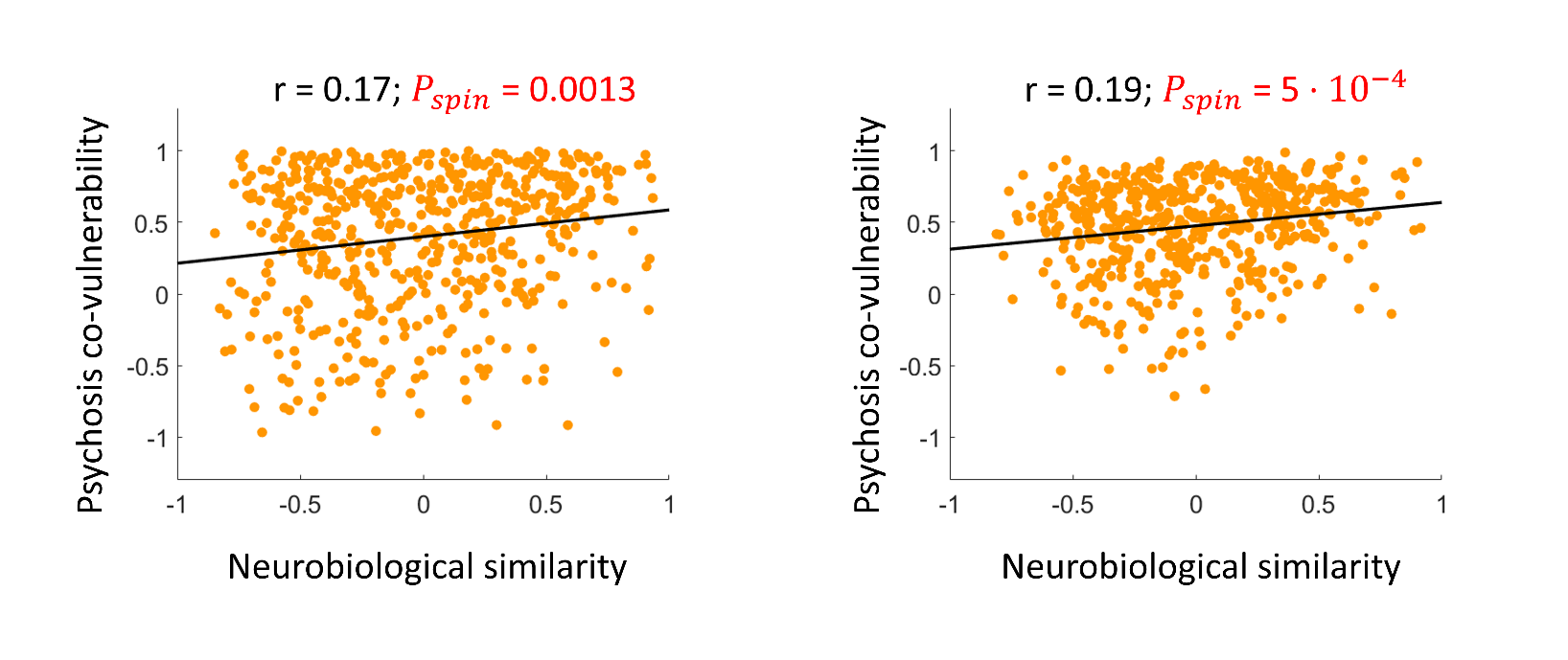

**Supplementary Fig. 21** Association between neurobiological similarity of the top-10 positive and top-10 negative most contributing features and structural co-vulnerability to psychosis across groups (SCZ and SAD-relatives, PE, FEP, and SCZ and SAD-chronic).

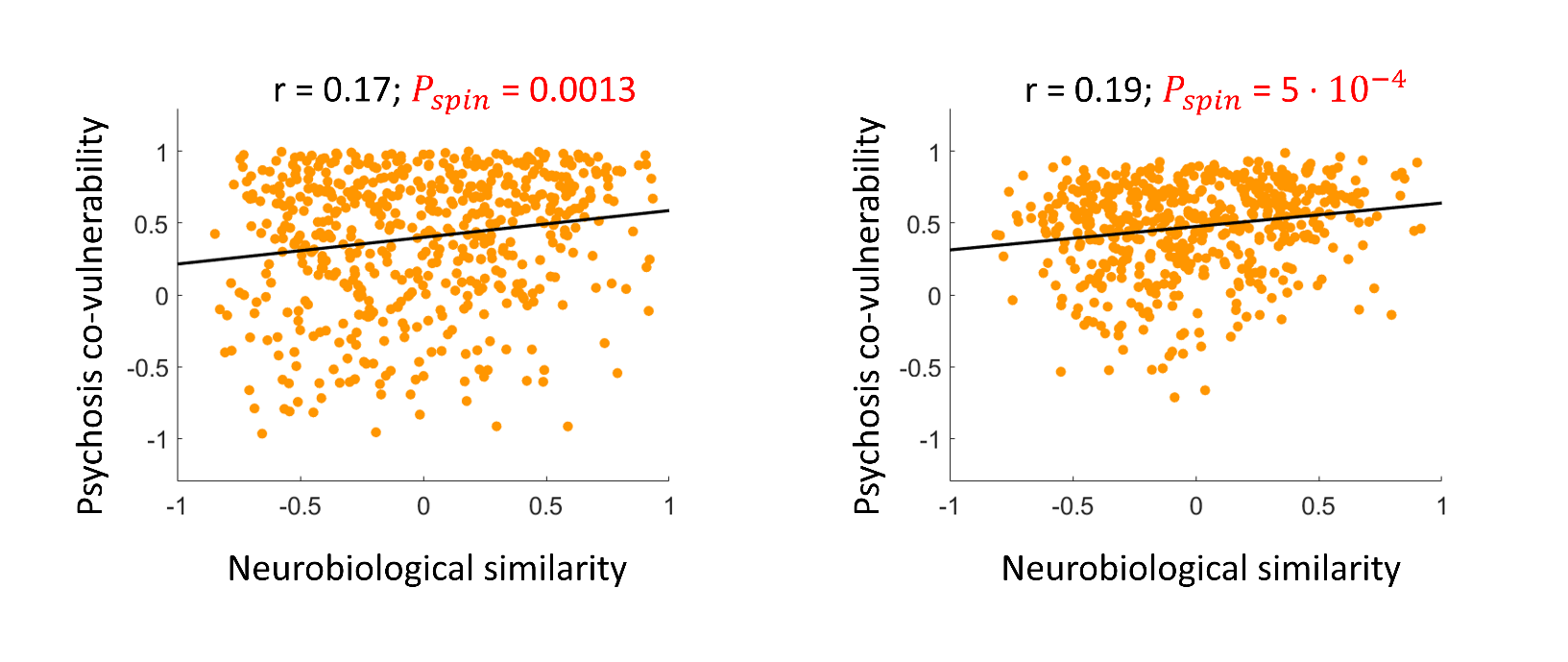

**Supplementary Fig. 22** Association between neurobiological similarity of the top-10 positive and top-10 negative most contributing features and structural co-vulnerability to psychosis for diagnoses (SCZ-relatives, SAD-relatives, PE-suspected, PE-definite, PE-clinical, FEP, SCZ, and SAD).

### Supplemental Tables

**Supplementary Table 1 Sample description for each diagnosis and its respective healthy control**

| **Group** | **Diagnosis** | **n (females)** | **Age (min, max)** | **Cohort (n; females)** |
| --- | --- | --- | --- | --- |
| SCZ and SAD-chronic | SCZ | 525 (148) | 37.63 ± 12.06 (10.19, 77.82) | ABCD (2; 1), ASRB (257; 76), BSNIP (100; 30), LA5c (48; 11), MCIC (104; 25), UKB (14; 5) |
|  | SAD | 62 (37) | 33.77 ± 10.78 (11.68, 57.06) | B-SNIP |
| FEP | FEP | 352 (138) | 31.43 ± 8.78 (18.78, 59.81) | PAFIP |
| PE | PE-suspected | 48 (33) | 21.55 ± 1.06 (20.78, 24.78) | ALSPAC |
|  | PE-definite | 73 (42) | 21.75 ± 1.59 (19.78, 24.78) | ALSPAC |
|  | PE-clinical | 36 (28) | 20.89 ± 0.92 (19.78, 23.78) | ALSPAC |
| SCZ and SAD-relatives | SCZ-relatives | 96 (68) | 43.75 ± 15.25 (11.68, 65.07) | B-SNIP |
|  | SAD-relatives | 64 (43) | 40.00 ± 16.20 (11.68, 65.07) | B-SNIP |
| Healthy Controls (HC) | HC | 38,232 (19,139) | 51.91 ± 23.68 (9.76, 81.82) | ABCD (8,762; 4,500), ASRB (177; 92), BSNIP (93; 59), LA5c (121; 57), MCIC (90; 28), UKB (28,989; 14,403) |
|  | HC | 195 (75) | 30.64 ± 7.66 (16.78, 52.80) | PAFIP |
|  | HC | 269 (154) | 22.30 ± 1.46 (19.78, 24.78) | ALSPAC |

PAFIP, *Programa de Atención a las Fases Iniciales de Psicosis*; ALSPAC, Avon Longitudinal Study of Parents and Children; B-SNIP, Bipolar & Schizophrenia Consortium for Parsing Intermediate Phenotypes; ABCD, Adolescent Brain and Cognitive Development; ASRB, Australian Schizophrenia Research Bank; LA5c, UCLA Consortium for Neuropsychiatric Phenomics LA5c Study; MCIC, Mental Illness and Neuroscience Discovery (MIND) Institute Clinical Imaging Consortium; UKB, UK Biobank.

**Supplementary Table 2 Neurobiological features included in PCA-CCA analyses and the neurobiological similarity matrix**

| **Abbreviation** | **Neurobiological feature** | **Type** |
| --- | --- | --- |
| 5-HT_1A_ | Serotonin receptor | Neurotransmitter |
| 5-HT1_B_ | Serotonin receptor | Neurotransmitter |
| 5-HT_2A_ | Serotonin receptor | Neurotransmitter |
| 5-HT_4_ | Serotonin receptor | Neurotransmitter |
| 5-HT_6_ | Serotonin receptor | Neurotransmitter |
| 5-HTT | Serotonin transporter | Neurotransmitter |
| H_3_ | Histamine receptor | Neurotransmitter |
| D_1_ | Dopamine receptor | Neurotransmitter |
| D_2_ | Dopamine receptor | Neurotransmitter |
| DAT | Dopamine transporter | Neurotransmitter |
| NET | Norepinephrine transporter | Neurotransmitter |
| α_4_β_2_ | Acetylcholine receptor | Neurotransmitter |
| M_1_ | Acetylcholine receptor | Neurotransmitter |
| VAChT | Acetylcholine transporter | Neurotransmitter |
| CB_1_ | Cannabinoid receptor | Neurotransmitter |
| MOR | Opioid receptor | Neurotransmitter |
| mGluR_5_ | Glutamate receptor | Neurotransmitter |
| NMDA | Glutamate receptor | Neurotransmitter |
| GABA | GABA receptor | Neurotransmitter |
| Astro | Astrocytes | Cell type |
| Endo | Endothelial cells | Cell type |
| Micro | Microglia | Cell type |
| Oligo | Oligodendrocytes | Cell type |
| OPC | Oligodendrocytes precursors | Cell type |
| Neuro-Ex | Excitatory neurons | Cell type |
| Neuro-In | Inhibitory neurons | Cell type |
| Layer I | Layer I | Layer thickness |
| Layer II | Layer II | Layer thickness |
| Layer III | Layer III | Layer thickness |
| Layer IV | Layer IV | Layer thickness |
| Layer V | Layer V | Layer thickness |
| Layer VI | Layer VI | Layer thickness |
| Myelin | Myelin | Microstructure |
| Thickness | Cortical thickness | Microstructure |
| Gene PC1 | Gene expression PC1 | Microstructure |
| Neurotransmitter PC1 | Neurotransmitter PC1 | Microstructure |
| Synapse density | Synapse density | Microstructure |
| Evolutionary exp. | Evolutionary expansion | Cortical expansion |
| Developmental exp. | Developmental expansion | Cortical expansion |
| Scaling PNC | Allometric scaling from Philadelphia Neurodevelopmental Cohort | Cortical expansion |
| Scaling NIH | Allometric scaling from National Institutes of Health | Cortical expansion |
| CBF | Cerebral blood flow | Metabolism |
| CBV | Cerebral blood volume | Metabolism |
| CMRO_2_ | Cerebral metabolic rate of oxygen | Metabolism |
| CMRGlu | Cerebral metabolic rate of glucose | Metabolism |
| Glycolytic index | Glycolytic index | Metabolism |
